# Assessing the accuracy of California county level COVID-19 hospitalization forecasts to inform public policy decision making

**DOI:** 10.1101/2022.11.08.22282086

**Authors:** Lauren A. White, Ryan McCorvie, David Crow, Seema Jain, Tomás M. León

**Affiliations:** California Department of Public Health, Richmond, CA, USA

**Keywords:** infectious disease modeling, forecasting, model evaluation, COVID-19, public health

## Abstract

**Background:** The COVID-19 pandemic has highlighted the role of infectious disease forecasting in informing public policy. However, significant barriers remain for effectively linking infectious disease forecasts to public health decision making, including a lack of model validation. Forecasting model performance and accuracy should be evaluated retrospectively to understand under which conditions models were reliable and could be improved in the future.

**Methods:** Using archived forecasts from the California Department of Public Health’s California COVID Assessment Tool (https://calcat.covid19.ca.gov/cacovidmodels/), we compared how well different forecasting models predicted COVID-19 hospitalization census across California counties and regions during periods of Alpha, Delta, and Omicron variant predominance.

**Results:** Based on mean absolute error estimates, forecasting models had variable performance across counties and through time. When accounting for model availability across counties and dates, some individual models performed consistently better than the ensemble model, but model rankings still differed across counties. Local transmission trends, variant prevalence, and county population size were informative predictors for determining which model performed best for a given county based on a random forest classification analysis. Overall, the ensemble model performed worse in less populous counties, in part because of fewer model contributors in these locations.

**Conclusions:** Ensemble model predictions could be improved by incorporating geographic heterogeneity in model coverage and performance. Consistency in model reporting and improved model validation can strengthen the role of infectious disease forecasting in real-time public health decision making.

## Background

In public health, forecasting has been used to predict infectious disease dynamics for a variety of diseases including influenza, dengue fever, Ebola virus disease, Zika fever, and most recently COVID-19, which has highlighted the importance of infectious disease modeling to help inform public health decision making (1). Nevertheless, significant barriers remain for effectively linking infectious disease forecasts with public health decision making including a lack of model standardization and validation, and difficulty in successfully communicating model complexity and uncertainty (2). Moreover, public health practitioners may need different outcomes or indicators than what forecast models provide (2,3).

In June 2020, as part of the COVID-19 response, the California Department of Public Health’s (CDPH) COVID Modeling Team launched the <u>California Communicable diseases Assessment Tool (CalCAT)</u> to compile available COVID-19 models, mostly from academic groups, to inform policy and public health action (4). CalCAT provides nowcasts (R-effective estimates), forecasts (short-term predictions for hospitalizations, ICU admissions, and deaths), and longer-range scenario models for a variety of COVID indicators at the state, regional, and county scales. Some contributors are national, forecasting for all states, while others focus only on California and may not be publicly available elsewhere (Table 1). The models on CalCAT have been used throughout the COVID-19 pandemic to evaluate current transmission trends and prospective hospital and intensive care unit capacity. This information combined with other evidence and policy considerations has helped to inform the implementation of stay-at-home orders and statewide mask mandates (e.g., reinstating a mask mandate during the emergence of Omicron/BA.1). In addition, models combined with other data streams were used to inform metrics for the Blueprint for a Safer Economy including the nation’s first health equity metric and to support planning for vaccine allocation and distribution (5).

**Table 1.** Constituent models providing county-level hospitalization census predictions that are archived on CalCAT and included in the analysis.

| **Model** | **Forecast update frequency** | **Forecast horizon** | **Methods/Approach** | **Documentation** |
| --- | --- | --- | --- | --- |
| Columbia | Weekly | Up to 6 weeks | County level metapopulation model | (6) |
| UCSF, COVID NearTerm | Daily | 2-4 weeks | Bootstrap-based method based on an autoregressive model | (7) |
| UCB LEMMA | Daily | Up to 4 weeks | SEIR compartmental model with parameters fit using case series data of COVID-19 hospital and ICU census, hospital admissions, deaths, cases and seroprevalence | (8) |
| CDPH Simple Growth | Daily | Up to 4 weeks | Assumes new cases grow exponentially according to the rate given by the latest ensemble R-effective. Assumes a fixed severity and average length of stay to generate hospitalizations | (4) |
| CalCAT Ensemble | Daily | Up to 4 weeks | The ensemble forecast takes the median of all the forecasts available on a given date and fits a smoothed spline to the trend. | (4) |
| CA Baseline | Daily | Up to 4 weeks | Retroactive 7-day rolling average mean of past hospitalization values | Methods |

| **Model** | **Forecast update frequency** | **Forecast horizon** | **Methods/Approach** | **Documentation** |
| --- | --- | --- | --- | --- |
| Columbia | Weekly | Up to 6 weeks | County level metapopulation model | (5) |
| UCSF, COVID NearTerm | Daily | 2-4 weeks | Bootstrap-based method based on an autoregressive model | (6) |
| UCB LEMMA | Daily | Up to 4 weeks | SEIR compartmental model with parameters fit using case series data of COVID-19 hospital and ICU census, hospital admissions, deaths, cases and seroprevalence | (7) |
| CDPH Simple Growth | Daily | Up to 4 weeks | Assumes new cases grow exponentially according to the rate given by the latest ensemble R-effective. Assumes a fixed severity and average length of stay to generate hospitalizations | (4) |
| CalCAT Ensemble | Daily | Up to 4 weeks | The ensemble forecast takes the median of all the forecasts available on a given date and fits a smoothed spline to the trend. | (4) |
| CA Baseline | Daily | Up to 4 weeks | Retroactive 7-day rolling average mean of past hospitalization values | Methods |

During the COVID-19 pandemic response, many California local health jurisdictions communicated the importance of forecasts focused on the relevant scale of decision making (e.g., county- vs. state-level forecasts) because there was significant geographic heterogeneity in COVID-19 outcomes at regional and local levels (9). A better understanding of how forecasting models have captured these geographical heterogeneities could help inform local public health decision making during future COVID-19 waves and enable local health jurisdictions to employ models judiciously given proven past performance. Lessons learned from COVID-19 forecasting efforts can also be applied to future modeling for other diseases including influenza.

We retrospectively evaluated archived forecasting predictions from CalCAT for models that consistently provided county-level hospitalization census predictions across a year long period from February 1, 2021 to February 1, 2022 (Table 1). Hereafter, we will use hospital census to refer to the number of patients currently hospitalized with confirmed COVID-19 for a given county and date. To explore the effects of COVID-19 variants on model performance within that period, we also compared forecasting model accuracy during three phases of the COVID-19 pandemic at the county and regional level in California (Figure 1 A-C) with different variant predominance: Alpha (April 22- June 1, 2021), Delta (June 21 - September 1, 2021), and Omicron (December 21, 2021 - February 1, 2022). These periods also differed in their hospitalization burden (Figure 1A) and epidemic growth rates (Figure 1C).

**Figure 1.**
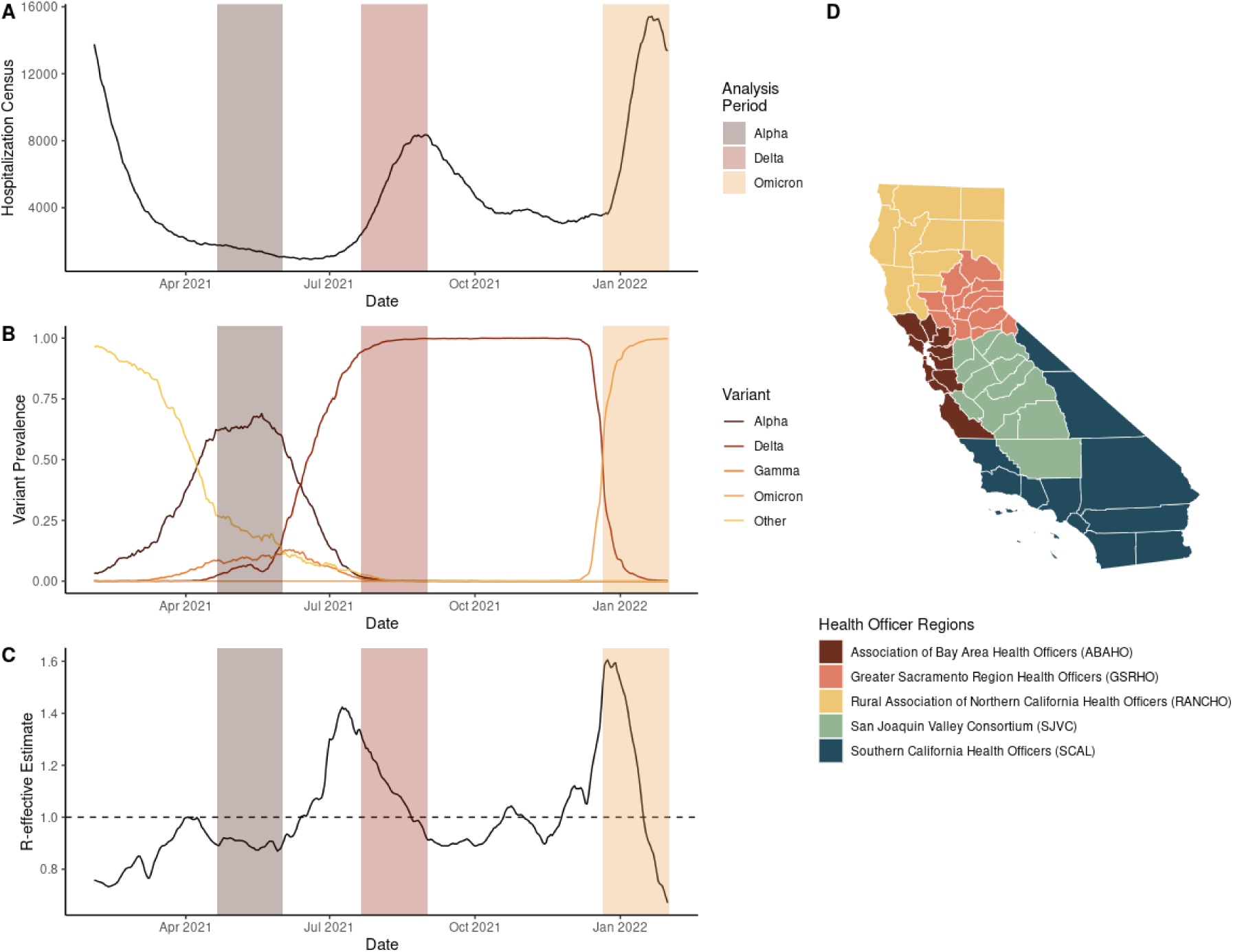
Time courses of (A) California COVID-19 hospitalization census, (B) variant prevalence, (C) statewide R-effective estimate, and (D) California health officer regions. The period displayed for panels A-C corresponds to the complete period of analysis February 1, 2021- February 1, 2022 used for the pairwise tournament and random forest analyses. Shaded regions for panels A:C correspond to the dates of analysis for the three variant predominant periods: Alpha, Delta, and Omicron.

## Methods

Multiple methods exist for measuring epidemic forecast accuracy including metrics that evaluate specific point estimates and uncertainty (10). When full predictive estimates are available, metrics like the logarithmic score or continuous ranked probability score (CRPS) provide context for probabilistic models’ predictions and uncertainty. When forecasts are provided in quantile or interval formats, the weighted interval score (WIS) is a potential alternative (10). Since not all models incorporated into CalCAT provided full predictive or interval estimates, or did so with different reporting standards, we focused on the median point estimates (50th percentile) from forecasting models for hospital census at the county scale. In addition to these models, we retroactively created a baseline California forecast that projected forward the 7-day rolling mean from the prior week. Each forecast has the following properties: (1) model *(m):* the organization or group issuing the forecast (Table 1); (2) location (*j*): the geographic location for which a forecast was issued (in this case, at the county-level): (3) publication date *(i):* the date that the forecast was displayed on CalCAT; and (4) target end date *(k):* the future forecast horizon date for which the prediction was made.

We utilized mean absolute error (MAE) and relative error at the 7-, 14-, and 21-day forecast horizons to evaluate the accuracy of these point estimates. To better compare across counties with different population sizes, we normalized both error types by the median hospital capacity of each county (14-day horizon results are highlighted in the main text; the remaining forecast horizons are provided in the Supporting Information). From the MAE scores, we computed a standardized ranking score for every forecasted observation relative to other models issuing a prediction for that same publication date and location (11). In addition, we also conducted pairwise tournaments of model performance to control for the frequency of model participation. Finally, using a classification regression approach, we explored which county-level epidemiological and socio-economic covariates could help explain the “winning” model for a given location and date based on the lowest MAE scores for a given forecast horizon.

County results are grouped by health officer regions, which are contiguous groupings of 58 counties used for health mandates in California (Figure 1D): Association of Bay Area Health Officers (ABAHO); Greater Sacramento Region Health Officers (GSRHO); Rural Association of Northern California Health Officers (RANCHO); San Joaquin Valley Consortium (SJVC); and Southern California (SCAL). Some counties do not have major hospitals and therefore lack forecasting predictions, actual numbers of hospitalizations, or both. For this reason, Alpine, Sierra, and Sutter Counties are not included in the analyses that follow.

### Mean absolute error

The raw mean absolute error (MAE) for each publication date *i* with associated target end dates *k* is calculated as: 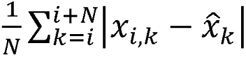 where *N* is the number of days into the future that the forecast is made, *x_i k_* is the prediction made on publication date *i* for target end date *k* and 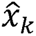 is the actual observed value for a given target end date (11). We then standardized the MAE by *h,* the median non-surge hospital capacity of a given county: 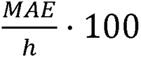

Median hospital capacity was chosen for standardization because the hospital capacity for facilities, and aggregated for counties, changes through time based on staffing and other factors. Note that not all model forecasts were available for all counties or all dates. A model only received an MAE score for a given publication date if it had predictions available for the target end dates of interest (e.g., to receive a 7-day MAE score, a model must have made predictions for 1-7 days ahead of the publication date). Here we used CA-state specific data (12) for post-hoc evaluation, whereas many model teams may be relying on other data sources (e.g., U.S. Department of Health and Human Services) for fitting or calibration.

### Relative error

The relative error for each publication date *(i)* with associated target end dates *k* is calculated as: 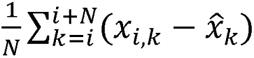 where *N* is the number of days into the future that the forecast is made, *x_i,k_* is the prediction made on publication date *i* for target end date *k* and 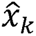 is the actual observed value for a given target end date (11). We then standardized the relative error by *h,* the median non-surge hospital capacity of a given county: 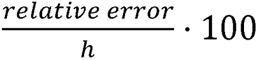

Therefore, a positive relative error corresponds to a model overestimating the hospital census, while a negative relative error corresponds to an underestimation.

### Standardized ranking score

For each publication date *i* and location *j, we* calculated a standardized rank for every available model *m* based on its associated MAE: 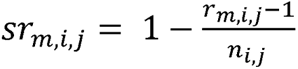 where *r_m,i,j_* is the ranking of the MAE of model *m* out of the *n* other models that made predictions for publication date *i* and location *j* (adapted slightly from (11)). Thus, for a given publication date *i* and location *j*, the highest possible standardized ranking score for any given model is 1 and the lowest is the inverse of the lowest possible ranking (l/*n_i,j_*). Models not participating for a given publication date *i* and location *j* receive a zero, and thus, are penalized for lack of coverage.

### Pairwise tournament

To conduct a pairwise tournament, we calculated a relative MAE for each pair of models *m* and 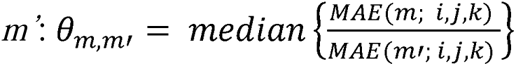 where *θ_m,m_*’is the median of the ratio of the simultaneously available MAE scores for model *m* to model *m’* with shared publication dates *i*, target end dates *k*, and locations *j* (13). Importantly, the common locations, publication dates, and observation dates may differ for each pair of models *m* and *m’.* This approach varies slightly from some previous examples, as the order of operations is scale then aggregate rather than aggregate then scale (11,14).

An overall performance score of a given model, *m* is then calculated as the geometric mean of all relative MAE scores: 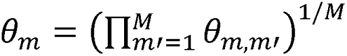 where*M* is the total number of all models available for comparison. At the county level for counties with smaller hospital capacities, there was a non-trivial probability of certain models achieving an MAE of zero, which leads to relative MAE scores of zero or infinity depending on the order of comparison. To eliminate these irregularities in the pairwise comparisons, we excluded counties with median non-surge hospital capacities ≤ 25 (i.e., Calaveras, Lassen, Mariposa, Modoc, Mono, San Benito, and Trinity).

### Random forest classification analysis

To explore whether the model with the lowest MAE score for a given location and observation date could be explained by county-specific epidemiological or socioeconomic factors, we conducted a random forest classification analysis. Random forest analysis is a recursive partitioning method that improves classification accuracy by synthesizing the predictions from many classification trees (15,16). The response variable (i.e., classification label) was the best performing model for a given county and date combination based on the lowest MAE score of the available models. We explored the covariates (i.e., features) of: progressive vaccination coverage at the county level, county-level R-effective, 7-day change in county-level R-effective, variant prevalence at the health officer region level (17), county population size, percent of county residents in poverty (2019), percent unemployment (2020), median income (2019), five-year average percentage completing college (university degree) (2015–2019), and 2013 Rural-Urban Continuum code. All socioeconomic variables were taken from U.S. Department of Agriculture Economic Research Service county-level data sets (18). For pre-processing, data were centered and scaled using the caret package (19). For model training and tuning, 70% of original data was used with K-fold validation (four-fold, repeated four times). The final accuracy of the random forest classification models were 61% with mtry = 7, 66% with mtry = 7, and 68% with mtry = 7 for 7, 14, and 21 day forecast horizons respectively.

### Data and code availability

The forecasts and R-effective values analyzed in this paper are available from CalCAT (4). California-specific hospitalization data is available on the California Open Data Portal (12). Because of reporting delays and backfilling, datasets used in the analysis may represent a snapshot of what was available at that point in time. All data and code used for analysis and figure generation is available in the public repository: https://doi.org/10.5281/zenodo.7851280. Analyses were performed in R (v 3.6.0) (20).

## Results

### Model performance varied across locations and under different periods of variant predominance

Model performance was heterogeneous across counties and during different periods of variant predominance (Figure 2A, 3A, 4A), in part reflecting that the number of models available for a given publication date and location varied through time; fewer models were available during the Omicron variant period and for less populous health officer regions such as RANCHO (Supplementary Figures 3, 11, 15). For example, in Trinity County – one of California’s least populous counties – the Simple Growth model had the lowest 14-day normalized MAE for most forecast publication dates during the Alpha and Omicron predominant periods (Figure 2A, 4A), whereas the Columbia model had the lowest 14-day normalized MAE during the Delta period (Figure 3A). In San Diego County, California’s second most populous county, the LEMMA model had the lowest 14-day normalized MAE during the Alpha period (Figure 2A), and the COVID NearTerm model had the lowest 14-day MAE for the most days during Delta and Omicron periods (Figure 3A & 4A). Overall, the Simple Growth model performed particularly well in the RANCHO region during the Omicron period as demonstrated by a lower 14-day MAE for many counties in that region (Figure 4A). The LEMMA model had the lowest 14-day MAE across many regions during the Omicron period on or after January 13, 2022 (Figure 4A). In general, the range of the relative error distributions increased with longer time horizons and during the Omicron period (Supplementary Figures 1-2). During the Omicron period, most relative error distributions were right skewed with median relative error values less than zero, indicating a tendency for underprediction, but a non-zero probability of sizeable overprediction (Supplementary Figure 1).

**Figure 2.**
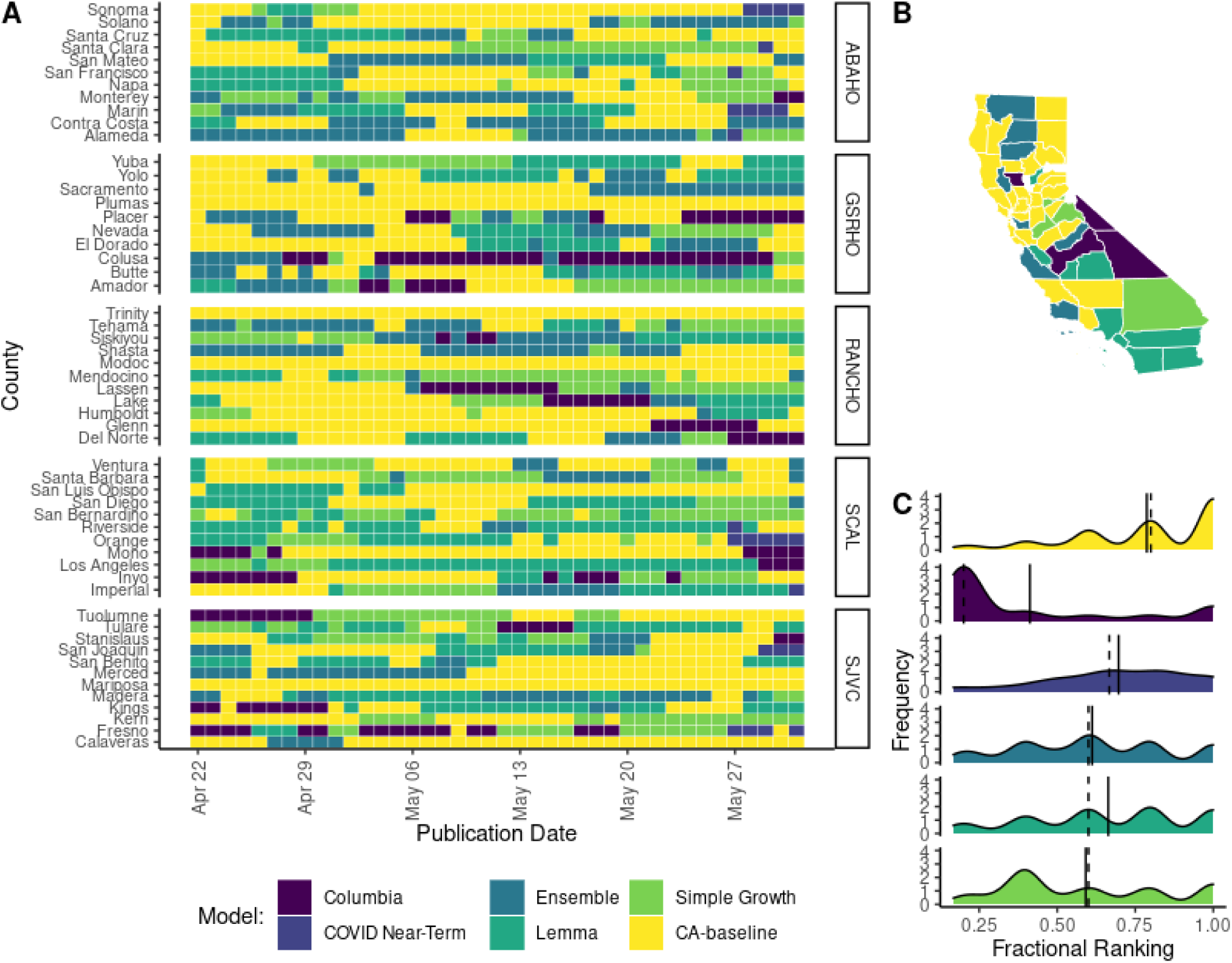
Forecasting accuracy results at the county level during the Alpha wave in California as measured by mean absolute error (MAE). (A) Heat map of the best daily performing model for a given prediction date as measured by 14-day MAE. Each cell in the heat map corresponds to a normalized MAE calculated for the day that a model forecast was published. Counties are grouped into panels by California health officer regions. (B) A summary map of California where the color of the county corresponds to the model with the highest sum of the standardized rank score for that period (*Σsr_m,i,j_).* Note that by using the summation of the standardized ranking score, models are penalized for lack of participation. (C) A density distribution of the standardized rank score (*sr_m,i,j_*) that depicts the median (dashed) and mean (solid) as vertical lines for each model distribution. A standardized rank score of one indicates that a model came in first relative to other participating models for a given date and location, values closer to zero indicate that a model had a lower ranking compared to other participating models, and a value of zero corresponds to no participation.

**Figure 3.**
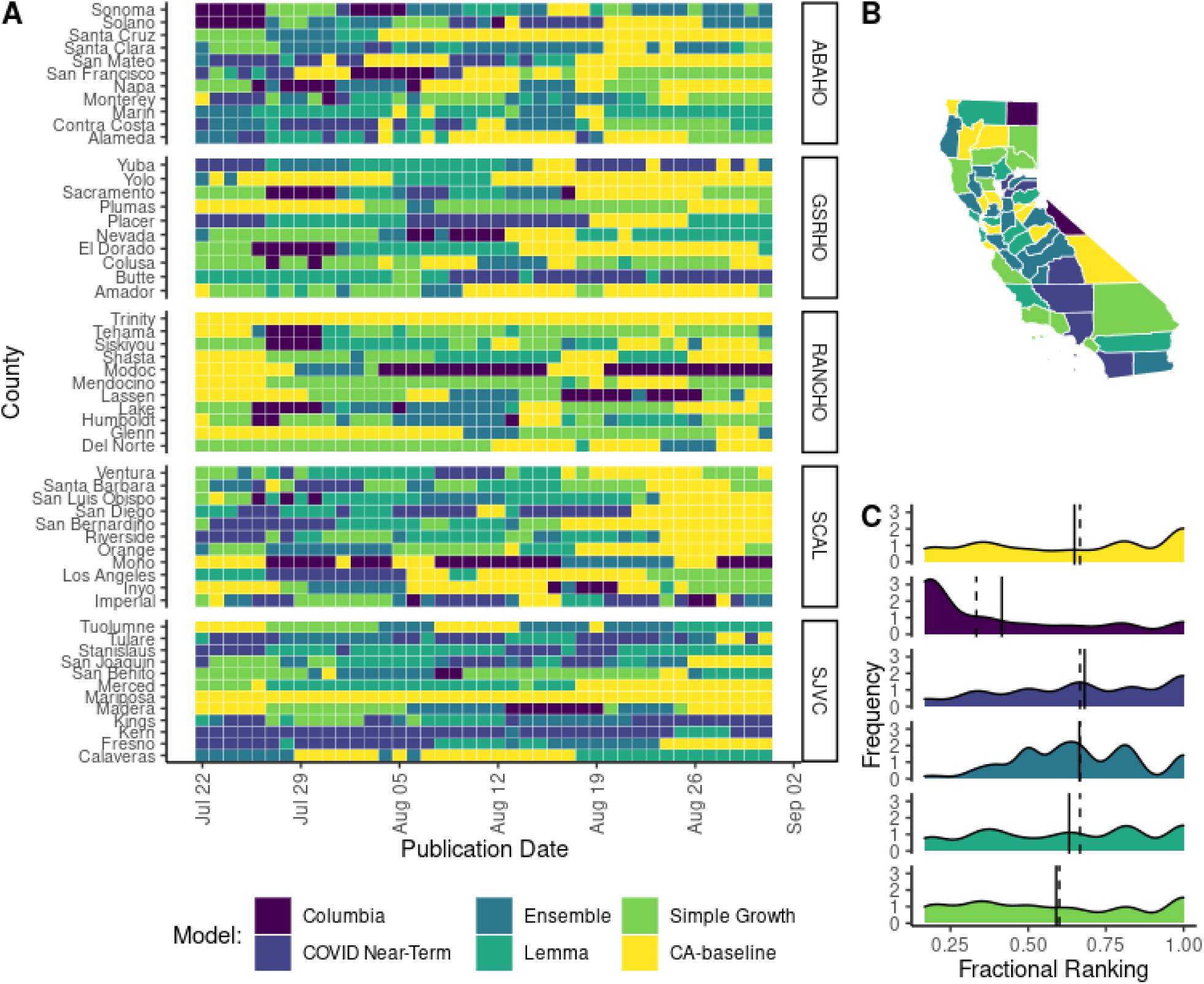
Forecasting accuracy results at the county level during the Delta wave in California as measured by mean absolute error (MAE). (A) Heat map of the best daily performing model for a given prediction date as measured by 14-day MAE. Each cell in the heat map corresponds to a standardized MAE calculated for the day that a model forecast was published. Counties are grouped into panels by California health officer regions. (B) A summary map of California where the color of the county corresponds to the model with the highest sum of the standardized rank score for that period (*Σsr_m,i,j_*). Note that by using the summation of the standardized ranking score models are penalized for lack of participation. (C) A density distribution of the standardized rank score *(sr_m,i,j_*) that depicts the median (dashed) and mean (solid) as vertical lines for each model distribution. A standardized rank score of one indicates that a model came in first relative to other participating models for a given date and location, values closer to zero indicate that a model had a lower ranking compared to other participating models, and a value of zero corresponds to no participation.

**Figure 4.**
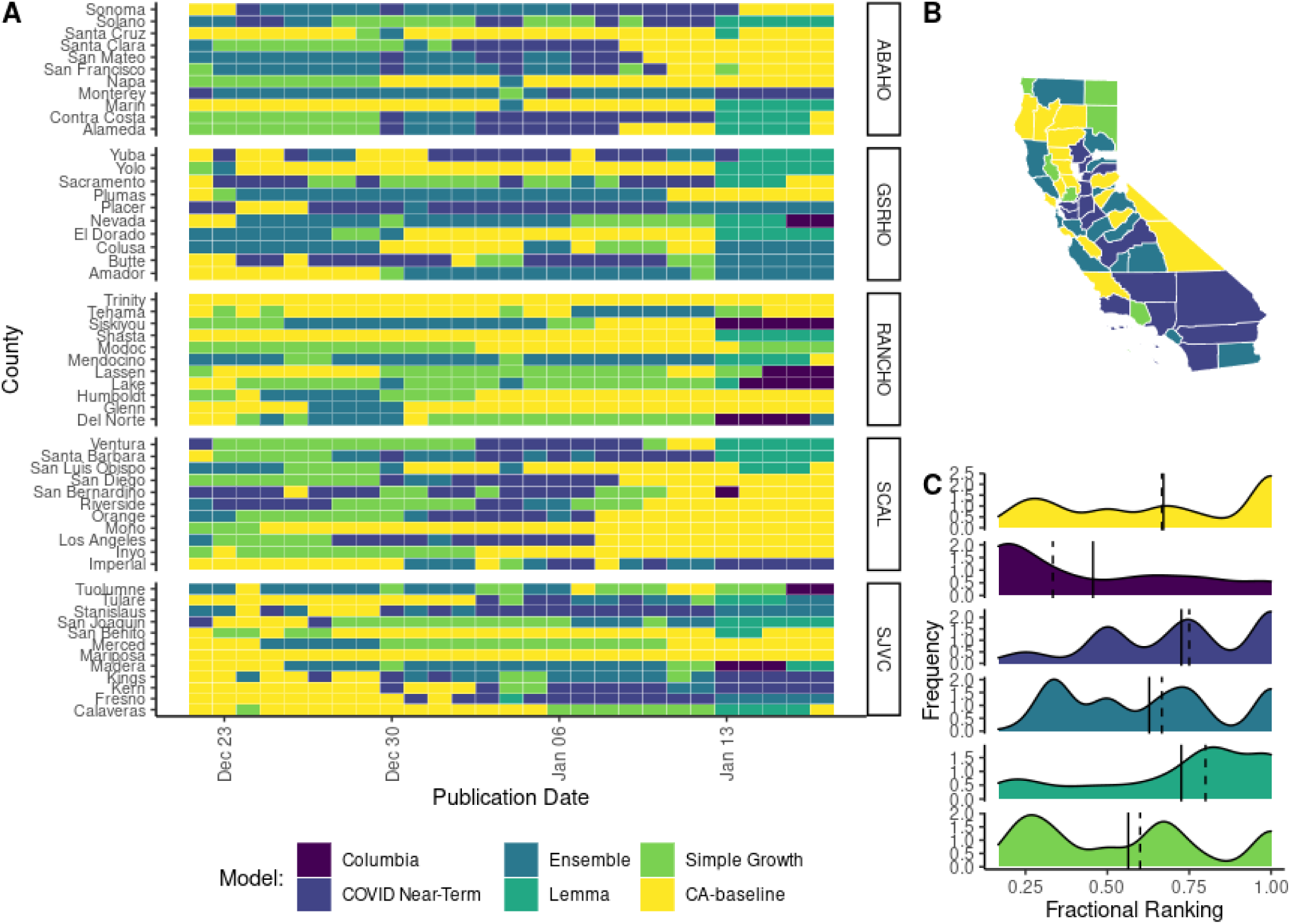
Forecasting accuracy results at the county level during the Omicron wave in California as measured by mean absolute error (MAE). (A) Heat map of the best daily performing model for a given prediction date as measured by 14-day MAE. Each cell in the heat map corresponds to a standardized MAE calculated for the day that a model forecast was published. Counties are grouped into panels by California health officer regions. (B) A summary map of California where the color of the county corresponds to the model with the highest sum of the standardized rank score for that period *Σsr_m,i,j_*). Note that by using the summation of the standardized ranking score models are penalized for lack of participation. (C) A density distribution of the standardized rank score (*sr_m,i,j_*) that depicts the median (dashed) and mean (solid) as vertical lines for each model distribution. A standardized rank score of one indicates that a model came in first relative to other participating models for a given date and location, values closer to zero indicate that a model had a lower ranking compared to other participating models, and a value of zero corresponds to no participation.

The sum of the standardized rank score (*Σsr_m,i,j_)* in each county, *j,* rewards both performance (model accuracy) and frequent participation (model coverage). During the Alpha period, the LEMMA model had the highest score in 20/55 counties, and the Ensemble model was a close second with the highest score in 17/55 counties (Figure 2B). During the Delta period, the Ensemble model had the highest score in 21/55 counties (Figure 3B). During the Omicron period, the Ensemble model had the highest score in 22/55 counties, and the Simple Growth model was a close second with 20/55 counties (Figure 4B).

The density distributions of standardized rank *(sr_m,i,j_)* allow for comparison of model performance while controlling for frequency of model participation (Figures 2C, 3C, 4C). Although COVID NearTerm did not have the highest sum of the standardized rank score in any counties, it had the highest median standardized rank score during the Alpha and Delta periods (Figures 2C, 3C). The LEMMA model had the highest median standardized. rank score during the Omicron period (Figure 4C). The same pattern of ranking was present for 7-day MAE (Supplementary Figures 4-6). For 21-day MAE, the COVID NearTerm model had the highest median standardized rank score during the Alpha and Omicron periods (Supplementary Figures 16 & 18), while the LEMMA model had the highest median rank scores during the Delta period (Supplementary Figure 17).

### When controlling for participation, some models outperformed the ensemble, but pairwise model rankings varied across counties

When matching across all locations and all observation dates, two models– COVID NearTerm and LEMMA– performed better in pairwise comparisons relative to the Ensemble model for 14-day MAE (Figure 5A & B). However, pairwise rankings were quite variable when disaggregated by county and also highlighted the differences in coverage and availability across locations for different models (Figure 5C). For example, although the Simple Growth model came fourth in the overall pairwise ranking (Figure 5A), it came first in twelve individual counties. Similarly, the Columbia model came last in the overall pairwise ranking (Figure 5A) and generally performed worse than average *(θ_m_ >* 1), but performed better than average *(θ_m_ <* 1) in Plumas and Inyo counties (Figure 5C).

**Figure 5.**
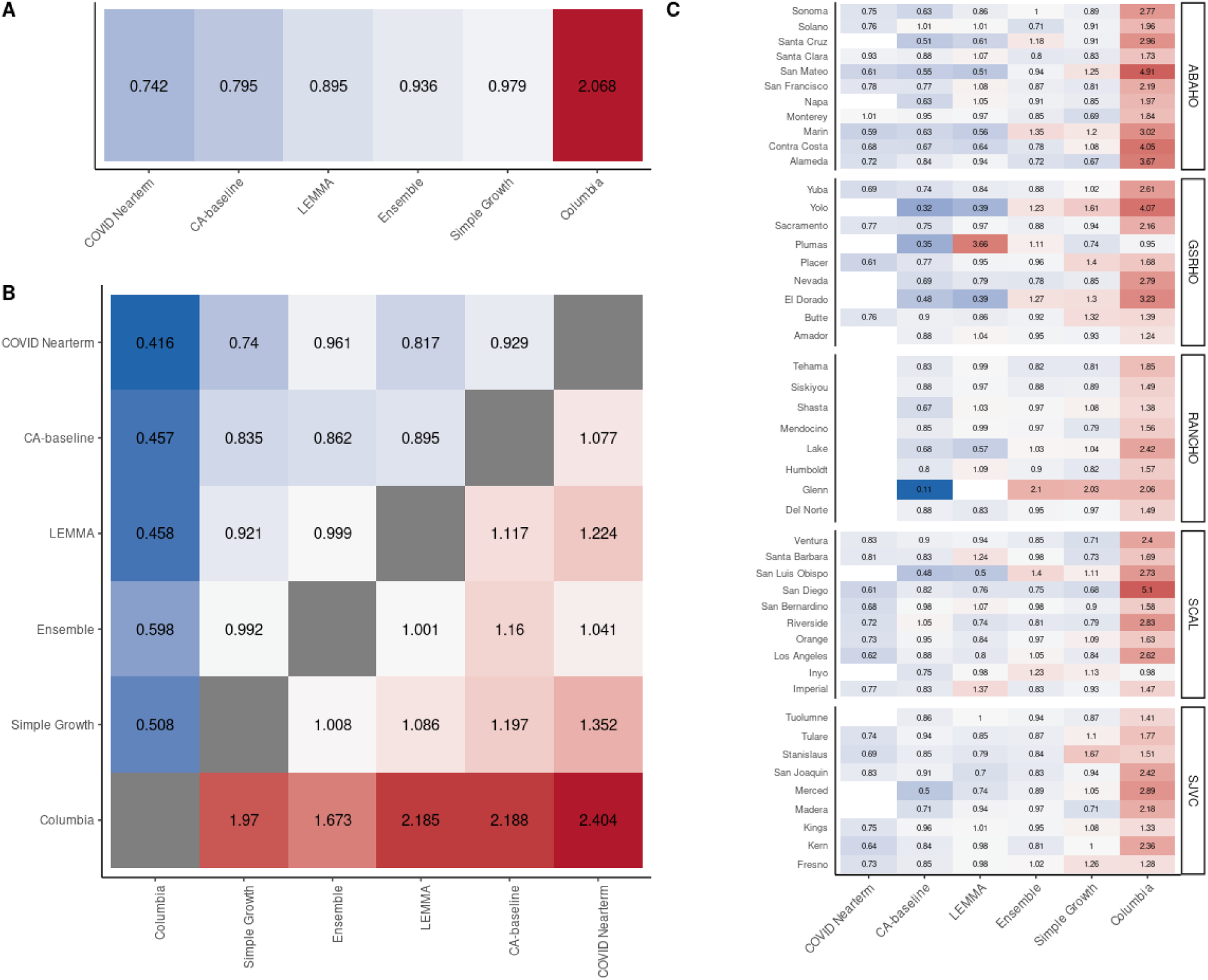
Pairwise tournament median rankings of models for the whole analysis period for 14-day MAE. **(A)** Overall median rankings (*θ_m_*) across all locations and observation dates. **(B)** Median pairwise rankings (*θ_m,m’_*) comparing each model *m* relative to every other model *m’.* The grid is symmetrical, so the ratio of model *m:* model *m’* is the inverse score of the ratio of model *m’:* model *m*. **(C)** Overall median rankings for all available observation dates disaggregated by county.

Overall pairwise rankings were robust to forecast horizon length for the complete analysis period (Supplementary Figures 5A & 17A). However, overall pairwise rankings were more unstable during specific periods of variant predominance, particularly for shorter forecast horizons (Supplementary Figures 6-8, 10-12, 18-20) and for county-specific rankings (Supplementary Figures 7C-9C, 12C-14C, 19C-21C).

### Epidemiological traits, county population size, and variant traits best predicted forecast “winners”

For the entire analysis period (February 1, 2021-February 1, 2022), time-varying vaccine coverage at the county-level, local transmission dynamics (R-effective and 7-day change in R-effective), county population size, and regional proportion of variants, were most important in predicting which model had the lowest MAE for a given county on a given publication date (Supplementary Figure 23). Other static socio-economic variables like income, percent unemployment, percent of residents with a university degree, and percent of residents in poverty were less important for predicting model outcomes. These variable importance rankings were robust to the forecast horizon used for MAE calculations (Supplementary Figure 23).

### Less populated counties have ensemble predictions with higher median MAE and more variable MAE

When comparing 14-day MAE normalized by hospital capacity, counties with smaller population sizes typically had a higher median MAE score and more variable MAE distributions compared to more populous counties (Supplementary Figure 24B). Based on a linear regression, the logarithmic of the normalized MAE score was negatively correlated with county population size (coefficient estimate: 1.7 10·10^−7^; p-value < 2·10^−16^). This relationship held true regardless of the forecast horizon used for MAE calculations (Supplementary Figure 24 A & C).

## Discussion

### Ensemble model could be improved by incorporating geographic heterogeneity in model coverage and performance

Echoing other analyses of COVID-19 forecast performance that have described a large variation in model accuracy by location (11,21), forecasting models performed differentially across California counties and regions and for different periods of variant predominance during the COVID pandemic (Figures 2-4, 5C, Supplementary Figures 4­6, 16-18). Moreover, location-specific features like local transmission dynamics or county population size helped explain model performance (Supplementary Figure 24). This geographic variation in model performance points to the importance of location-specific model evaluation in order for local health jurisdictions to best employ forecasts for public health decision making.

In general, combining multiple models into ensembles allows for better performance (11,22–25). However, in this case, COVID NearTerm and LEMMA consistently outperformed the Ensemble model when controlling for frequency of participation (Figures 2-4C, Figure 5), although pairwise ranking scores remained variable at the county level (Figure 5C). The higher performance of individual models over the Ensemble model combined with the variability in performance at the county-level suggests that the Ensemble model does not have to be applied uniformly across all locations; public health decision making could benefit from model selection and ensemble weighting that reflects location-specific past performance as well as local transmission trends (26).

### Lower forecast coverage in less populated counties weakens evidence-based decision making

One interesting question from a public health decision making context is whether model coverage (i.e., frequent issuing of forecasts across all potential locations) and model accuracy should be weighed equally when establishing the criteria for a “winning” forecast. In this analysis, there was typically a mismatch between raw model performance based on availability as measured by the sum of standardized ranking (Figures 2-4) and model performance when controlling for participation via pairwise tournaments (Figure 5). In part, this disagreement reflects that not all models provided estimates for all counties, especially for less populous regions or counties (Supplementary Figures 1, 9, 13). For example, the COVID NearTerm model ranked first in the pairwise ranking but provided no coverage for any counties in the less populous RANCHO region (Figure 5C). In contrast, the Ensemble model came first in the majority of counties during the Delta and Omicron periods as measured by sum of the standardized rank score for that period (*Σsr_m,i,j_)* (Figures 3B, 4B), but was generally outperformed in pairwise ranking evaluations both overall and for individual counties (Figure 5). Although the Ensemble model in less populous counties exhibited a higher median normalized MAE and a more variable normalized MAE regardless of the forecast horizon (Supplementary Figure 22), this observation may be a direct result of calculating MAE from median point estimates rather than accounting for forecast uncertainty, since stochastic effects likely contribute more significantly to the forecast predictions for counties with smaller population sizes.

While maximizing model accuracy is important, a forecast cannot add value if it is not available for decision making. As county-level contributors are lost to attrition, ensemble estimates may further decrease in accuracy or may not be possible in these less populous counties. Policy and public health decision makers should evaluate what investments or innovations in modeling are needed to improve results for underserved counties with lower forecast coverage. In addition, decision makers could seek to incentivize the best-performing models to serve smaller counties that neither have the resources to do this work in-house nor have academic partners readily available.

The lack of coverage in smaller counties also points to the inherent complexities of interpreting in hospitalization burden—since hospitalizations are typically recorded via hospital location rather than patient residency (27). As others have suggested, forecasting at the geographic unit of hospital referral networks could be another solution to low model coverage in less populous counties (27).

### Continuity of contributors, forecast structure, and documentation helps real-time public health decision making and post-hoc analysis

The overall continuity of forecasting contributions has proved challenging for post-hoc evaluation. Although CalCAT has had roughly ten unique forecast contributors through time, many of these groups have ceased contributing as the COVID-19 landscape has increased in complexity (e.g., emerging variants, prior immunity, boosters). Although less relevant to forecasting hospitalizations, changes in case ascertainment and testing practices make retrospective analyses more challenging. Interruptions to forecast continuity can also limit post-hoc evaluation. For example, some modeling groups paused forecasts in order to reset or recalibrate for new variants like Omicron.

While initiatives like the COVID-19 Forecast Hub have worked to standardize forecast output and reporting (11), one additional challenge for this analysis was that the reporting across external forecast contributors differed. For example, across three of the externally contributed forecasts they all produced interval estimates at different cutoff points: COVID Nearterm (10, 20, 30, 40, 50, 60, 70, 80, and 90 percentiles), Columbia (2.5, 25, 50, 75 and 97.5 percentiles), and LEMMA (5, 50, 95 percentiles). This discrepancy precluded the use of more robust measures like CRPS or WIS and means that that our results are much more sensitive to the median point estimates (10). Changing repository structures, file nomenclature, and data formatting can also disrupt the archiving process necessary for ensemble generation and subsequent post-hoc review. This analysis is a snapshot of what was available on CalCAT—and therefore to the general public and public health decision makers—and may not entirely reflect what model contributors would intend to be their contributing forecast at all times. The CalCAT team updated data and results iteratively as often as possible, but not all model changes were announced. As the COVID-19 pandemic necessitated rapid changes in data reporting and data infrastructure, other information technology issues may have introduced unintended errors.

The current classification regression analysis in this manuscript does not include model-specific traits. In order to truly evaluate whether underlying model traits and assumptions help to explain performance for specific locations through time, it would be necessary to have a larger number of forecasting contributors and consistent metadata on both the changes in model construction and the timing of those changes. Therefore, another potential area of documentation might include not just existing model assumptions and structure but how those characteristics have changed over time. This analysis may be easier to do at a state or national scale, where more model contributors are available, and reporting is better standardized through initiatives like the Forecast and Scenario Hubs.

Reporting and communicating infectious disease forecasting results, with all their inherent uncertainty and complexity, remain areas for improvement and growth for public health departments and their academic and industry collaborators to support evidence-based public health policy planning and decision making. Importantly, forecasting models may also serve as a communication tool to influence behavior change by the general public. One phenomenon not explored in this analysis is the potential for forecasts to alter human behavior, and subsequently model accuracy.

## Conclusions

Major progress in infectious disease forecasting has been made during the COVID-19 pandemic, while ongoing challenges, such as those around data and communication, have persisted. We retrospectively investigated hospitalization census forecast model performance at the county level in the state of California. Model performance and ranking varied through space and time and by metric, highlighting the difficulty of making blanket recommendations for which models to use for individual counties, including an ensemble approach. Calibrating based on past model performance may help improve ensemble forecast generation, and counties may benefit by considering which individual model contributors have historically served them the best. Going forward, closer collaboration between forecasters, researchers, and policymakers may create positive feedback loops that inform the ongoing COVID-19 response and other future public health action.

## List of abbreviations

(ABAHO): Association of Bay Area Health Officers
(CalCAT): California Communicable diseases Assessment Tool
(GSRHO): Greater Sacramento Region Health Officers
(MAE): Mean absolute error
(RANCHO): Rural Association of Northern California Health Officers
(SJVC): San Joaquin Valley Consortium
(SCAL): Southern California

## Declarations

### Ethics approval and consent to participate

Not applicable

## Consent for publication

Not applicable

## Availability of data and materials

The forecasts and R-effective values analyzed in this paper are available from CalCAT (4). California-specific hospitalization data is available on the California Open Data Portal (12). Because of reporting delays and backfilling, datasets used in the analysis may represent a snapshot of what was available at that point in time. All data and code used for analysis and figure generation is available in the public repository: https://github.com/whit1951/CA_COVID_Forecasting_Accuracy.

## Competing interests

The authors declare that they have no competing interests.

## Funding

This work was supported by the California Department of Public Health. The findings and conclusions in this article are those of the authors and do not necessarily represent the views or opinions of the California Department of Public Health or the California Health and Human Services Agency. This work was funded by Centers for Disease Control and Prevention, Epidemiology and Laboratory Capacity for Infectious Diseases, Cooperative Agreement Number 6 NU50CK000539.

## Authors’ contributions

LW and TL designed research and wrote the paper. LW performed the research and analyzed data. RM and DC contributed new analytic tools. RM, DC, and SJ revised/edited the manuscript. SJ supervised the project. All authors read and approved the final manuscript.

## Data Availability

All data and code used for analysis are available online at: https://doi.org/10.5281/zenodo.7851280

https://github.com/whit1951/CA_COVID_Forecasting_Accuracy

https://calcat.covid19.ca.gov/cacovidmodels/

https://data.ca.gov/group/covid-19

https://www.ers.usda.gov/data-products/county-level-data-sets/

https://doi.org/10.5281/zenodo.7851280

## Acknowledgments

The authors thank the CMU Delphi Group including Ryan Tibshurani and Daniel J. McDonald, MIDAS members, and the COVID-19 Forecasting Hub for discussion and feedback. The authors also thank Californian local health jurisdictions and members of the CDPH Modeling and Advanced Analytics team including Chris Hoover, Mugdha Thakur, Natalie Linton, Phoebe Lu, Sindhu Ravuri, and Sophie Zhu for conversations and insights that improved these analyses.

## Notes

### Competing Interest Statement

The authors have declared no competing interest.

### Author Declarations

Source data were openly available before the initiation of the study and located on the CA Open Data Portal for COVID-19 hospitalizations (https://data.ca.gov/dataset/covid-19-hospital-data1/resource/8f989799-b959-46ca-b3c5-0e67e95b584e) and COVID-19 vaccination coverage (https://data.ca.gov/dataset/covid-19-vaccine-progress-dashboard-data). Forecast data including hospitalization forecasts and R-effective estimates are available on the California Communicable Diseases Assessment Tool: https://calcat.covid19.ca.gov/cacovidmodels/

### Summary of Updates

- adding a column highlighting the forecasting horizon for each model to Table 1 - providing additional methodological details on the random forest classification analysis - clarifying the choice to use median point estimates and providing additional caveats for interpreting those results - explaining our choice of the median hospital capacity for normalization - provided context of the choice of definition for relative error - providing additional context for interpreting the standardized ranking score - adding additional caveats in the discussion of the relationship between mean absolute error and county populations size - providing some examples of public health decisions informed by models displayed on CalCAT

## References

1. Bertozzi AL, Franco E, Mohler G, Short MB, Sledge D. The challenges of modeling and forecasting the spread of COVID-19. Proc Natl Acad Sci. 2020 Jul 21;117(29):16732–8.

2. Lutz CS, Huynh MP, Schroeder M, Anyatonwu S, Dahlgren FS, Danyluk G, et al. Applying infectious disease forecasting to public health: a path forward using influenza forecasting examples. BMC Public Health. 2019 Dec;19(1):1659.

3. Doms C, Kramer SC, Shaman J. Assessing the Use of Influenza Forecasts and Epidemiological Modeling in Public Health Decision Making in the United States. Sci Rep. 2018 Dec;8(1):12406.

4. California Department of Public Health. California COVID Assessment Tool [Internet]. 2022. Available from: https://calcat.covid19.gov/cacovidmodels/

5. California Department of Public Health. Blueprint for a Safer Economy [Internet]. 2021 [cited 2023 Mar 6]. Available from: https://www.cdph.ca.gov/Programs/CID/DCDC/Pages/COVID-19/COVID19CountyMonitoringOverview.aspx

6. Pei S, Shaman J. Initial Simulation of SARS-CoV2 Spread and Intervention Effects in the Continental US [Internet]. Epidemiology; 2020 [cited 2022 Mar 22]. Available from: http://medrxiv.org/lookup/doi/10.1101/2020.03.21.20040303

7. Olshen AB, Garcia A, Kapphahn KI, Weng Y, Wesson PD, Rutherford GW, et al. COVIDNearTerm: A Simple Method to Forecast COVID-19 Hospitalizations [Internet]. Infectious Diseases (except HIV/AIDS); 2021 Oct [cited 2022 Mar 22]. Available from: http://medrxiv.org/lookup/doi/10.1101/2021.10.08.21264785

8. Schwab J, Peterson M. Local Epidemic Modeling for Management and Action (LEMMA) [Internet]. 2021. Available from: https://localepi.github.io/LEMMA/index.html

9. California State Government. California’s commitment to health equity [Internet]. California for All. 2022 [cited 2022 May 3]. Available from: https://covid19.ca.gov/equity/

10. Bracher J, Ray EL, Gneiting T, Reich NG. Evaluating epidemic forecasts in an interval format. Pitzer VE, editor. PLOS Comput Biol. 2021 Feb 12;17(2):e1008618.

11. Cramer EY, Ray EL, Lopez VK, Bracher J, Brennen A, Castro Rivadeneira AJ, et al. Evaluation of individual and ensemble probabilistic forecasts of COVID-19 mortality in the United States. Proc Natl Acad Sci U S A. 2022 Apr 12;119(15):e2113561119.

12. California Department of Public Health. COVID-19 Hospital Data [Internet]. California Open Data Portal. Available from: https://data.ca.gov/group/covid-19

13. Green A, Hu A, Jahja M, Ventura V, Wasserman L, Tibshirani R, et al. CMU Delphi Covid-19 Forecasts [Internet]. 2021. Available from: https://github.com/cmu-delphi/covid-19-forecast/tree/develop#delphi-forecasting-efforts

14. Bracher J. Evaluating probabilistic COVID19 forecasts under partial missingness: A pairwise comparison approach [Internet]. COVID-19 Forecast Hub; 2020 Oct 27. Available from: https://covid19forecasthub.org/talks/2020-10-27-Bracher_Pairwise_Comparisons.pdf

15. Breiman L. Random Forests. Mach Learn. 2001 Oct 1;45(1):5–32.

16. Cutler DR, Edwards Jr. TC, Beard KH, Cutler A, Hess KT, Gibson J, et al. Random Forests for Classification in Ecology. Ecology. 2007;88(11):2783–92.

17. California Department of Public Health. Variants [Internet]. https://covid19.ca.gov/variants/. Available from: https://covid19.ca.gov/variants/

18. U.S. Department of Agriculture Economic Research Service. County-level Data Sets [Internet]. Data Products. 2021 [cited 2022 Mar 23]. Available from: ers.usda.gov/data-products/county-level-data-sets/

19. Kuhn M. caret: Classification and Regression Training [Internet]. 2021. Available from: https://github.com/topepo/caret/

20. R Core Team. R: A language and environment for statistical computing [Internet]. Vienna, Austria: R Foundation for Statistical Computing; 2021. Available from: https://www.R-project.org/

21. Reich NG, Tibshirani RJ, Ray EL, Rosenfeld R. On the predictability of COVID-19 [Internet]. 2021 [cited 2022 Jun 3]. Available from: https://forecasters.org/blog/2021/09/28/on-the-predictability-of-covid-19/

22. Reich NG, McGowan CJ, Yamana TK, Tushar A, Ray EL, Osthus D, et al. Accuracy of real-time multi-model ensemble forecasts for seasonal influenza in the U.S. Pitzer VE, editor. PLOS Comput Biol. 2019 Nov 22;15(11):e1007486.

23. Johansson MA, Apfeldorf KM, Dobson S, Devita J, Buczak AL, Baugher B, et al. An open challenge to advance probabilistic forecasting for dengue epidemics. Proc Natl Acad Sci. 2019 Nov 26;116(48):24268–74.

24. Viboud C, Sun K, Gaffey R, Ajelli M, Fumanelli L, Merler S, et al. The RAPIDD ebola forecasting challenge: Synthesis and lessons learnt. Epidemics. 2018 Mar;22:13–21.

25. The Influenza Forecasting Working Group, McGowan CJ, Biggerstaff M, Johansson M, Apfeldorf KM, Ben-Nun M, et al. Collaborative efforts to forecast seasonal influenza in the United States, 2015–2016. Sci Rep. 2019 Dec;9(1):683.

26. Ray EL, Reich NG. Prediction of infectious disease epidemics via weighted density ensembles. Viboud C, editor. PLOS Comput Biol. 2018 Feb 20;14(2):e1005910.

27. Rosenfeld R, Tibshirani RJ. Epidemic tracking and forecasting: Lessons learned from a tumultuous year. Proc Natl Acad Sci U S A. 2021 Dec 21;118(51):e2111456118.

