## Supporting Information for "Assessing the accuracy of California county level COVID-19 hospitalization forecasts to inform public policy decision making"

### Relative Error

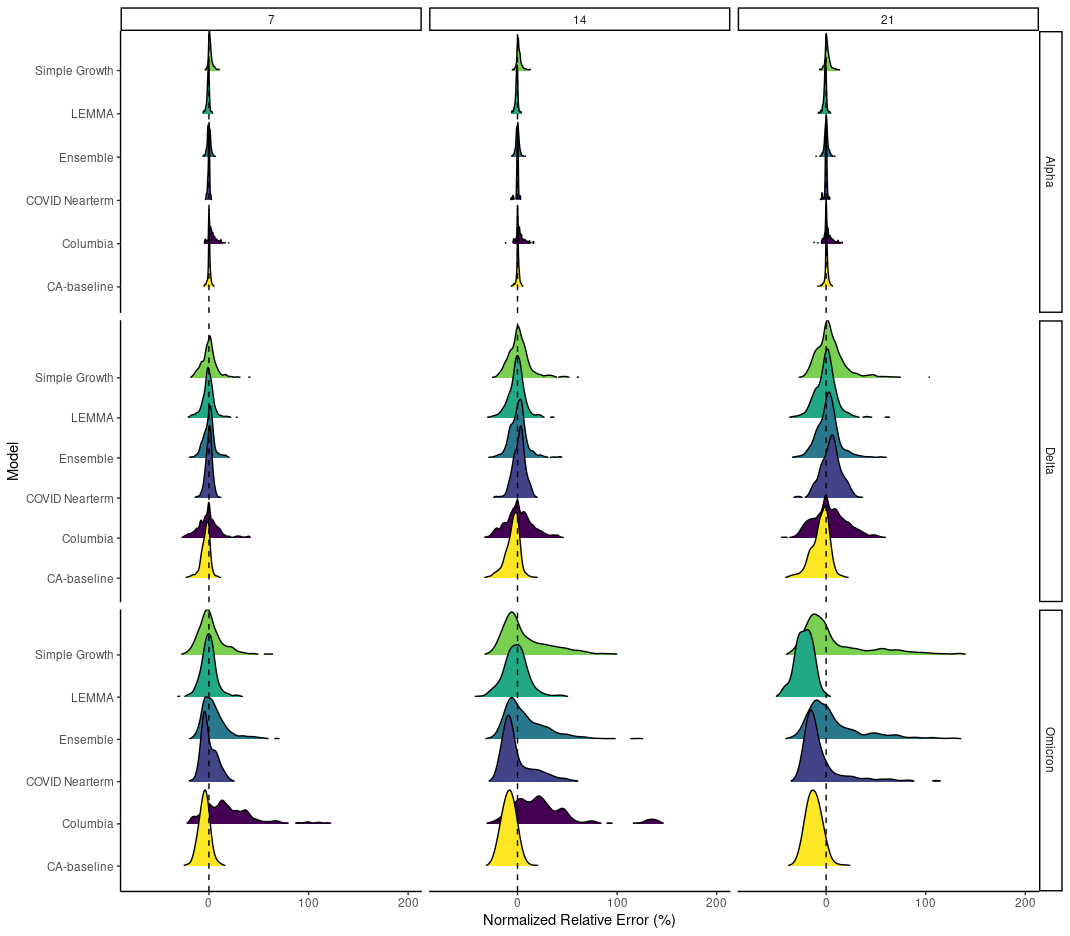

**Supplementary Figure 1. Relative error distributions across all counties for different forecast horizons (columns) and across different variant periods (rows) for the entire analysis period (February 1, 2021-Feburary 1, 2022).**

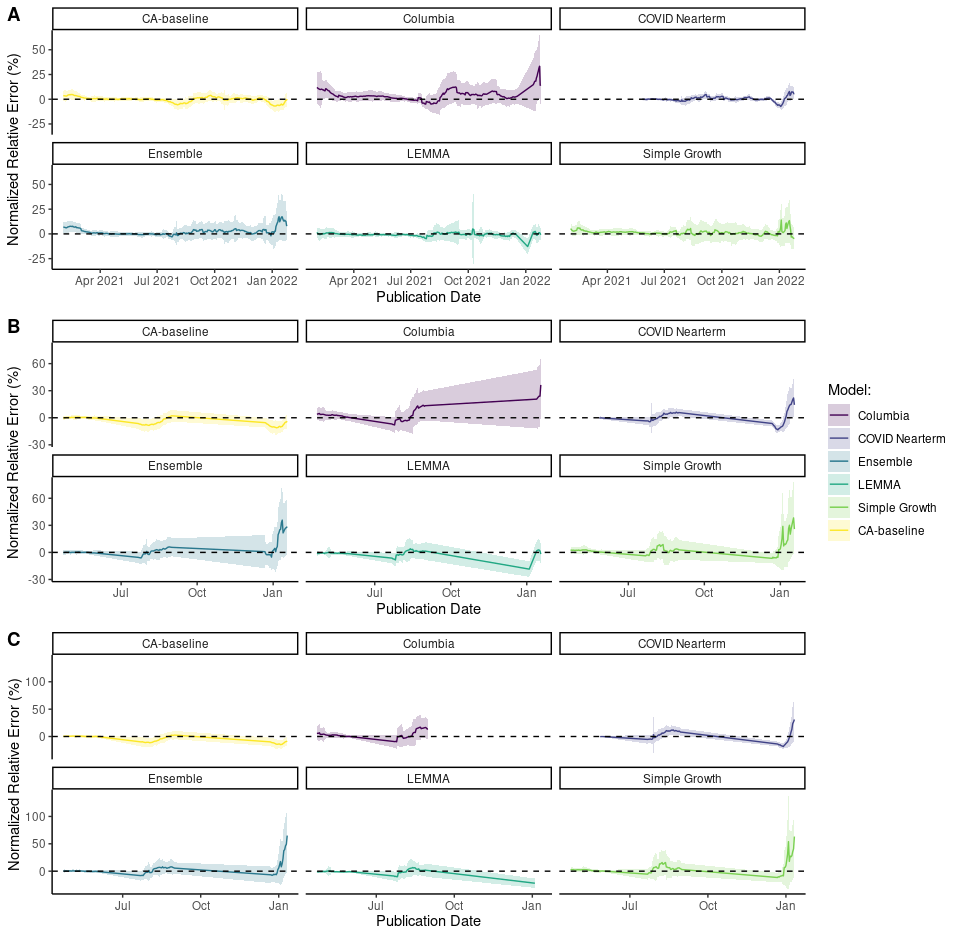

**Supplementary Figure 2. Normalized relative error distributions through the time for (A) 7-day, (B) 14-day, and (C) 21-day forecast horizons.**

### 7-day MAE

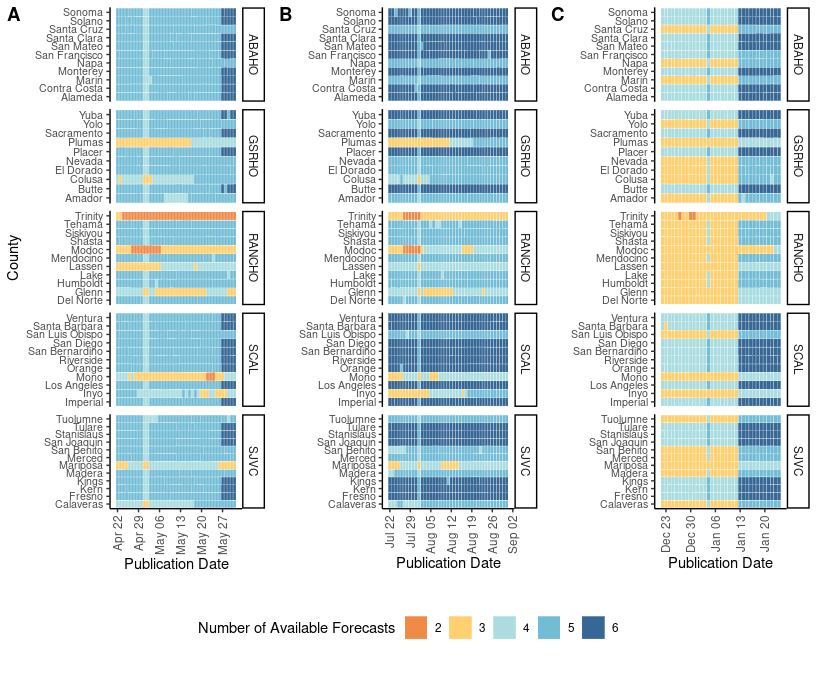

**Supplementary Figure 3. Number of available forecasts for a given location and date for the (A) Alpha, (B) Delta, and (C) Omicron variant periods for 14-day MAE.**

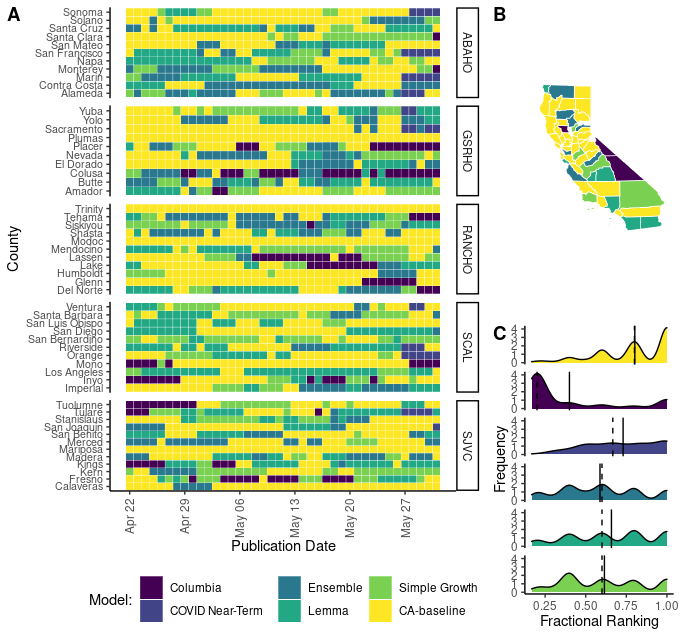

**Supplementary Figure 4. Forecasting accuracy results at the county level during the Alpha wave in California as measured by mean absolute error (MAE).** **(A)** Heat map of the best daily performing model for a given prediction date as measured by 7-day MAE. Each cell in the heat map corresponds to a standardized mean absolute error calculated for the day that a model forecast was published. Counties are grouped into panels by California health officer regions. **(B)** A summary map of California where the color of the county corresponds to the model with the highest sum of the standardized rank score for that period $({\Sigma sr}_{m,i,j})$. Note that by using the summation of the standardized ranking score models are penalized for lack of participation. **(C)** A density distribution of the standardized rank score $({sr}_{m,i,j})$ that depicts the median (dashed) and mean (solid) as vertical lines.

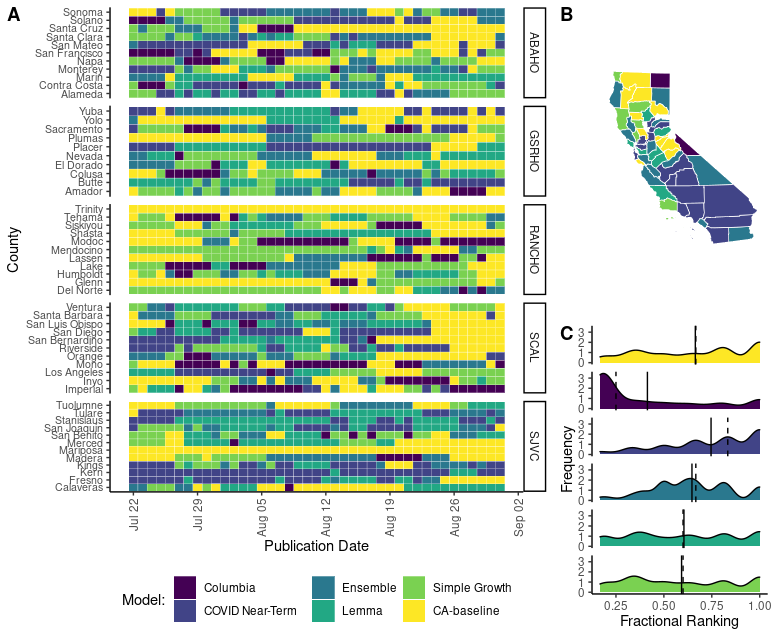

**Supplementary Figure 5. Forecasting accuracy results at the county level during the Delta wave in California as measured by mean absolute error (MAE).** **(A)** Heat map of the best daily performing model for a given prediction date as measured by 14 day MAE. Each cell in the heat map corresponds to a standardized mean absolute error calculated for the day that a model forecast was published. Counties are grouped into panels by California health officer regions. **(B)** A summary map of California where the color of the county corresponds to the model with the highest sum of the standardized rank score for that period $({\Sigma sr}_{m,i,j})$. Note that by using the summation of the standardized ranking score models are penalized for lack of participation. **(C)** A density distribution of the standardized rank score $({sr}_{m,i,j})$ that depicts the median (dashed) and mean (solid) as vertical lines.

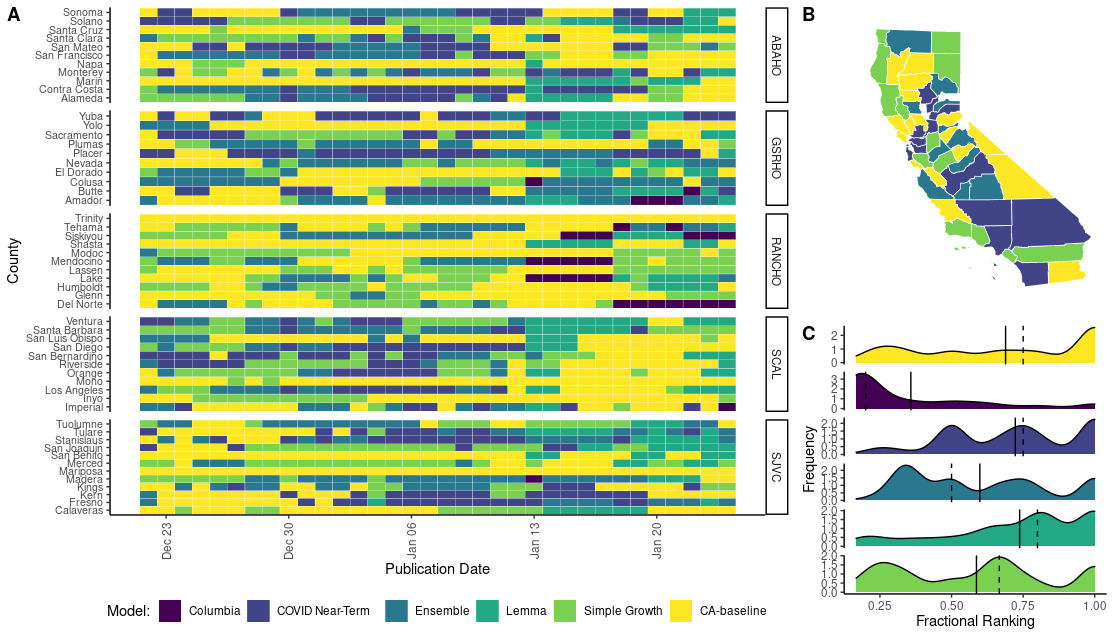

**Supplementary Figure 6. Forecasting accuracy results at the county level during the Omicron wave in California as measured by mean absolute error (MAE). (A)** Heat map of the best daily performing model for a given prediction date as measured by 7 day MAE. Each cell in the heat map corresponds to a standardized mean absolute error calculated for the day that a model forecast was published. Counties are grouped into panels by California health officer regions. **(B)** A summary map of California where the color of the county corresponds to the model with the highest sum of the standardized rank score for that period $({\Sigma sr}_{m,i,j})$. Note that by using the summation of the standardized ranking score models are penalized for lack of participation. **(C)** A density distribution of the standardized rank score $({sr}_{m,i,j})$ that depicts the median (dashed) and mean (solid) as vertical lines.

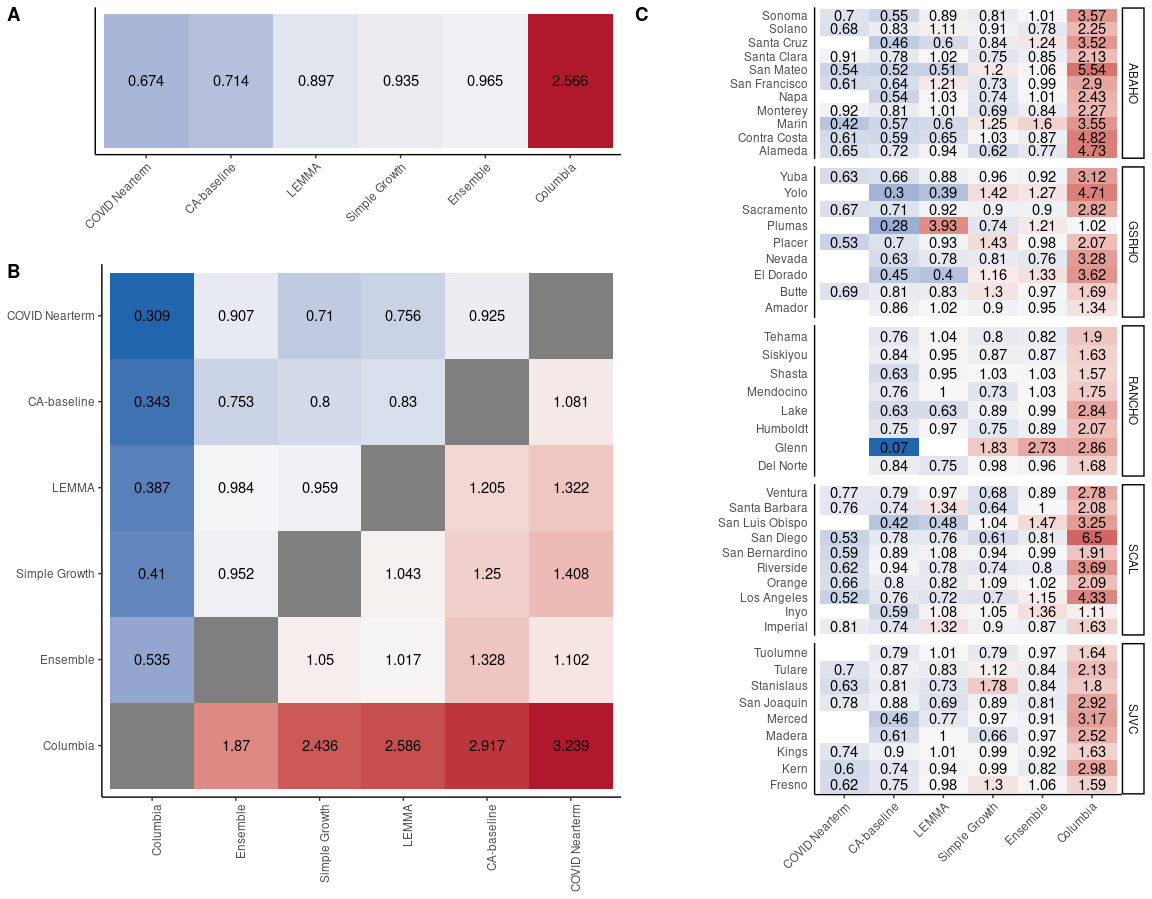

**Supplementary Figure 7. Pairwise tournament median rankings of models for the entire analysis period (January 1, 2021- February 1, 2022) for 7-day MAE.** **(A)** Overall median ranking across all locations and observation dates. **(B)** Median pairwise ranking comparing each model *m* to all available models *M*. Grid is symmetrical, so Model 1: Model 2 = 1/(Model 2:Model 1). **(C)** Overall median rankings for all available observation dates disaggregated by county.

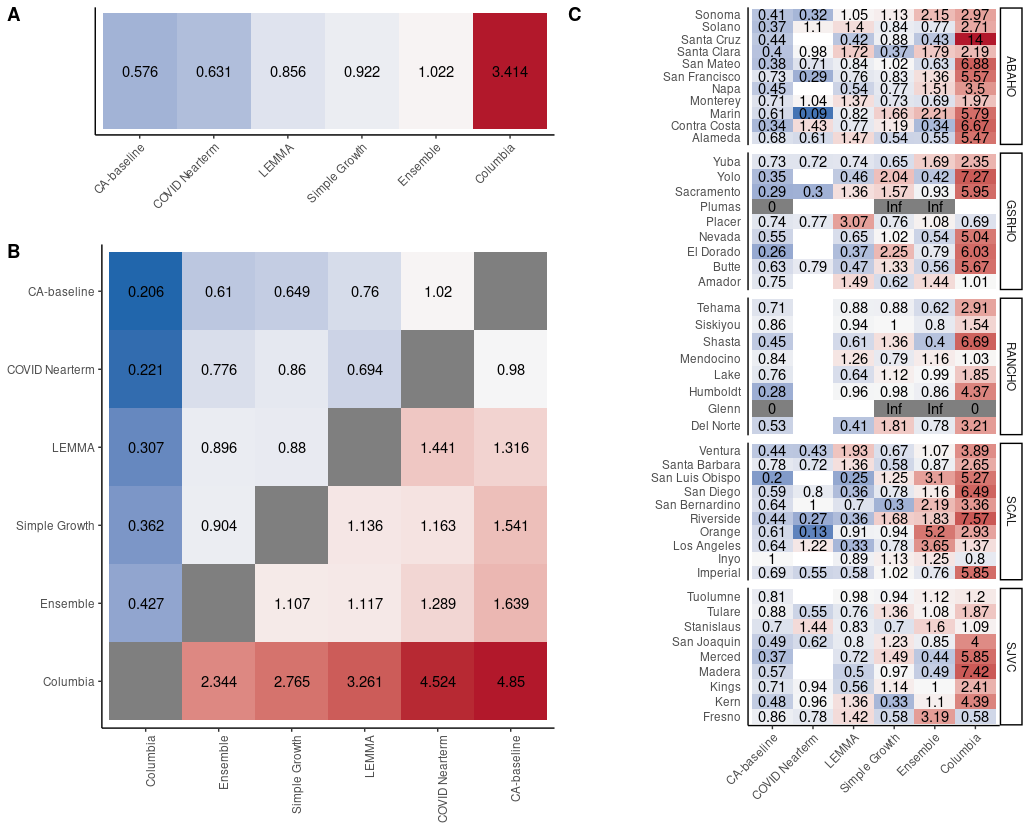

**Supplementary Figure 8. Pairwise tournament median rankings of models for the Alpha variant period for 7-day MAE. (A)** Overall median ranking across all locations and observation dates. **(B)** Median pairwise ranking comparing each model *m* to all available models *M*. Grid is symmetrical, so Model 1: Model 2 = 1/(Model 2:Model 1). **(C)** Overall median rankings for all available observation dates disaggregated by county.

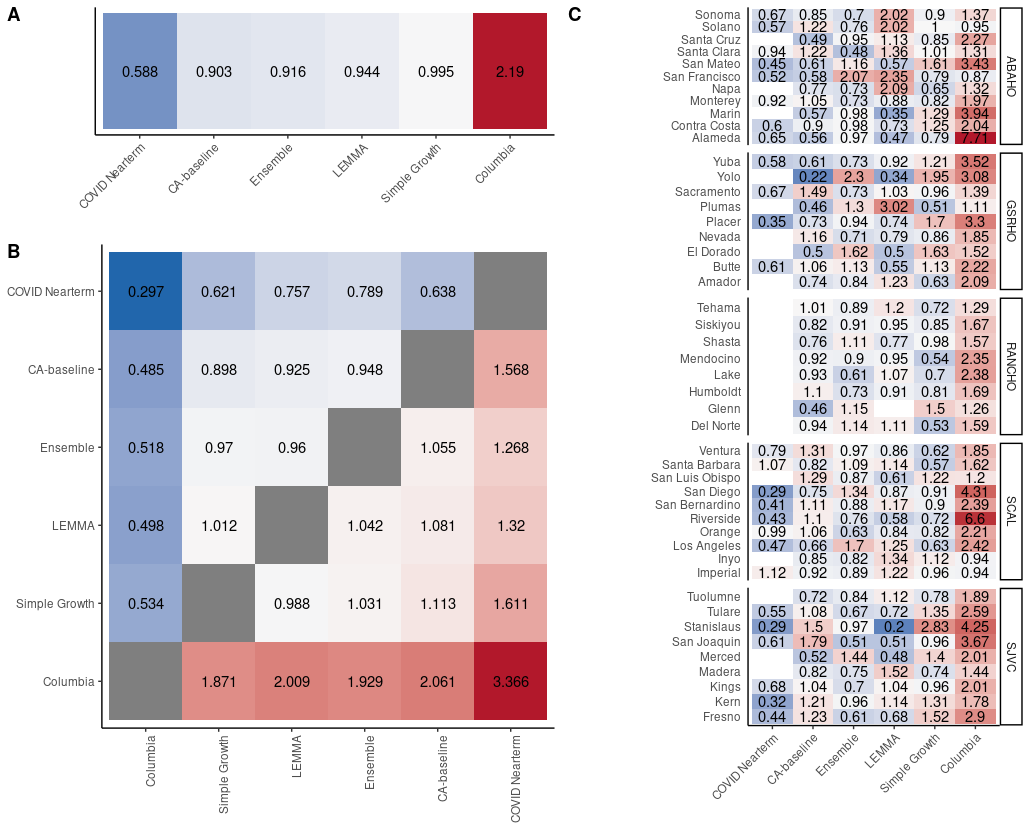

**Supplementary Figure 9. Pairwise tournament median rankings of models for the Delta variant period for 7-day MAE.** **(A)** Overall median ranking across all locations and observation dates. **(B)** Median pairwise ranking comparing each model *m* to all available models *M*. Grid is symmetrical, so Model 1: Model 2 = 1/(Model 2:Model 1). **(C)** Overall median rankings for all available observation dates disaggregated by county.

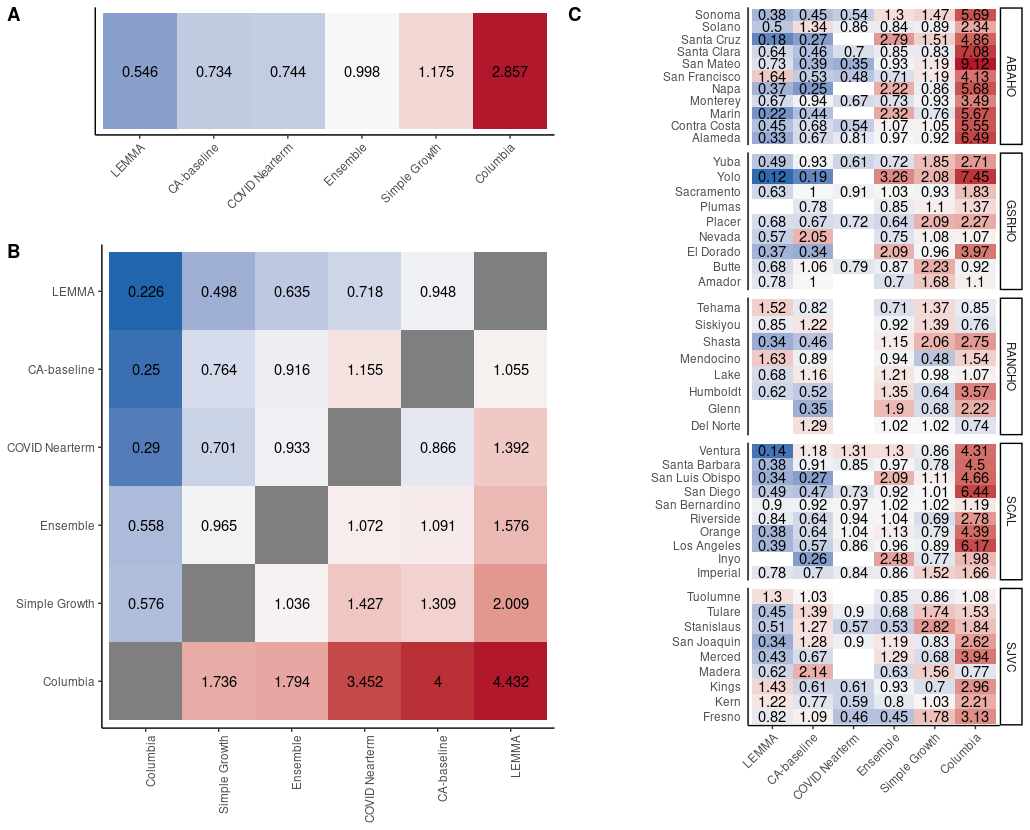

**Supplementary Figure 10.** **Pairwise tournament median rankings of models for the Omicron variant period for 7-day MAE.** **(A)** Overall median ranking across all locations and observation dates. **(B)** Median pairwise ranking comparing each model *m* to all available models *M*. Grid is symmetrical, so Model 1: Model 2 = 1/(Model 2:Model 1). **(C)** Overall median rankings for all available observation dates disaggregated by county.

### 14-day MAE

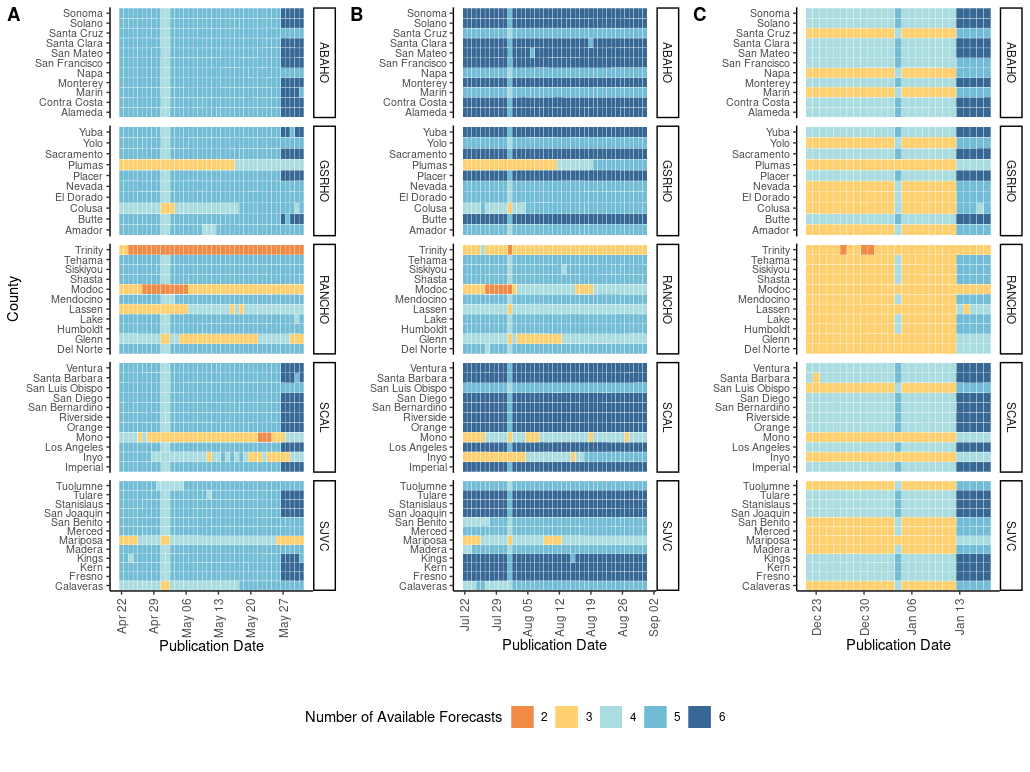

**Supplementary Figure 11. Number of available forecasts for a given location and date for the (A) Alpha, (B) Delta, and (C) Omicron variant periods for 14-day MAE.**

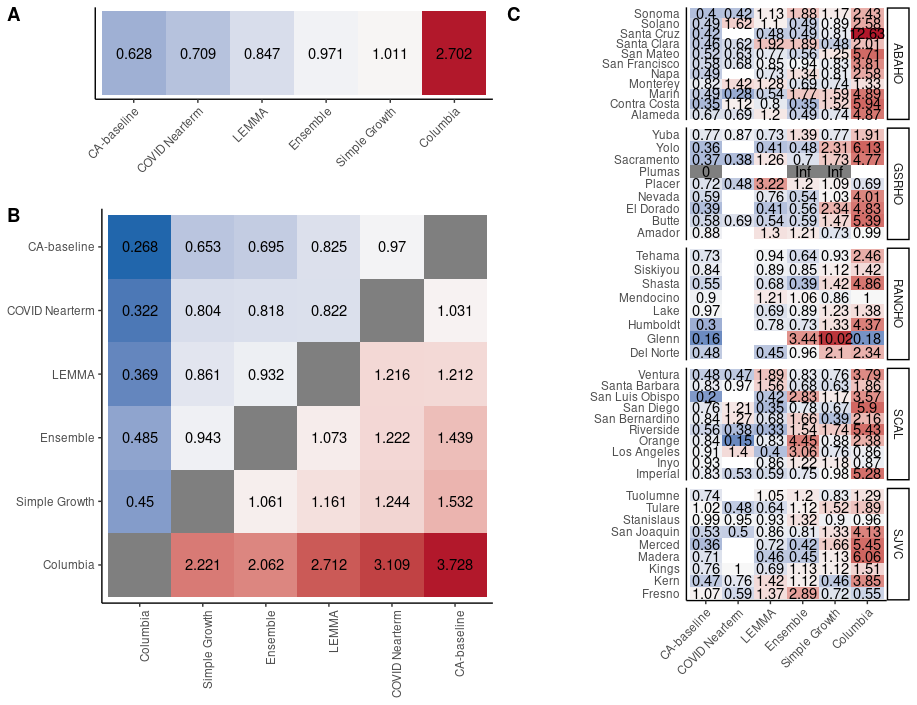

**Supplementary Figure 12.** **Pairwise tournament median rankings of models during the Alpha analysis period for 14-day MAE.** **(A)** Overall median ranking across all locations and observation dates. **(B)** Median pairwise ranking comparing each model *m* to all available models *M*. Grid is symmetrical, so Model 1: Model 2 = 1/(Model 2:Model 1). **(C)** Overall median rankings for all available observation dates disaggregated by county.

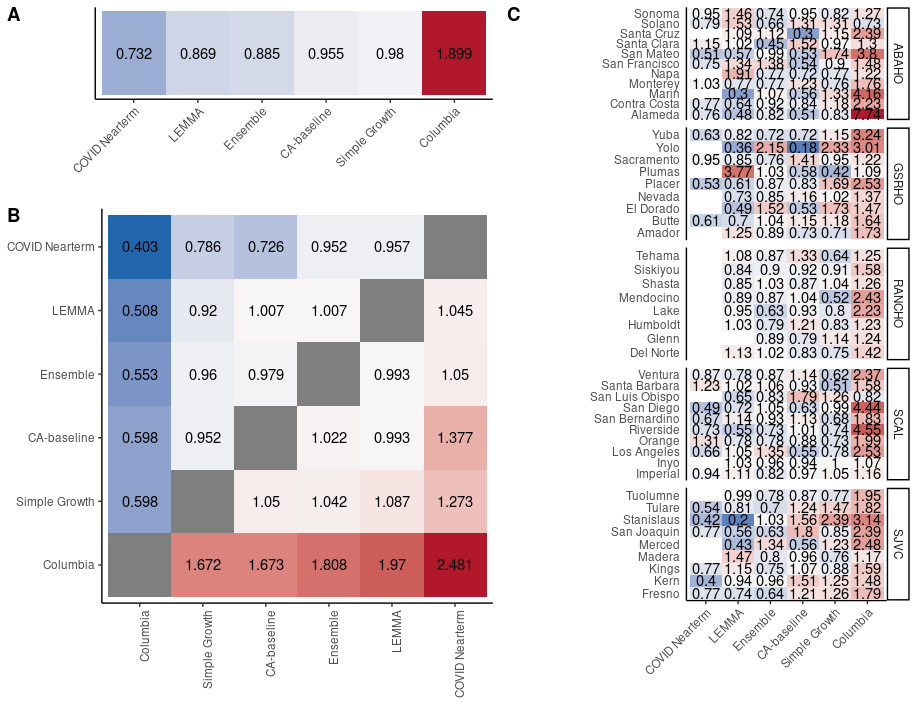
 **Supplementary Figure 13.** **Pairwise tournament median rankings of models during the Delta analysis period for 14-day MAE. (A)** Overall median ranking across all locations and observation dates. **(B)** Median pairwise ranking comparing each model *m* to all available models *M*. Grid is symmetrical, so Model 1: Model 2 = 1/(Model 2:Model 1). **(C)** Overall median rankings for all available observation dates disaggregated by county.

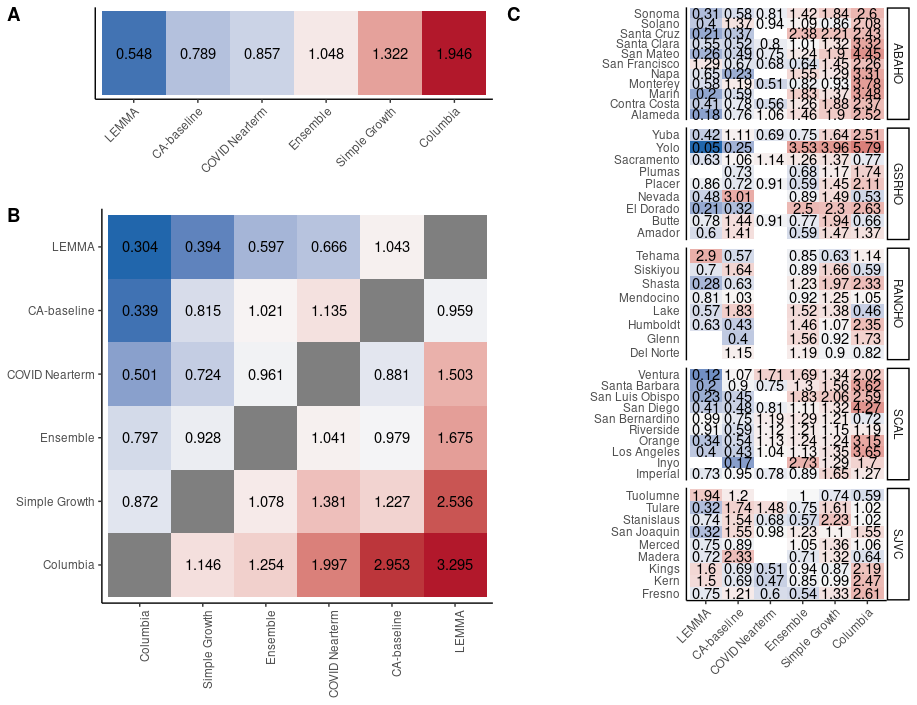

**Supplementary Figure 14.** **Pairwise tournament median rankings of models during the Omicron analysis period for 14-day MAE. (A)** Overall median ranking across all locations and observation dates. **(B)** Median pairwise ranking comparing each model *m* to all available models *M*. Grid is symmetrical, so Model 1: Model 2 = 1/(Model 2:Model 1). **(C)** Overall median rankings for all available observation dates disaggregated by county.

### 21-day MAE

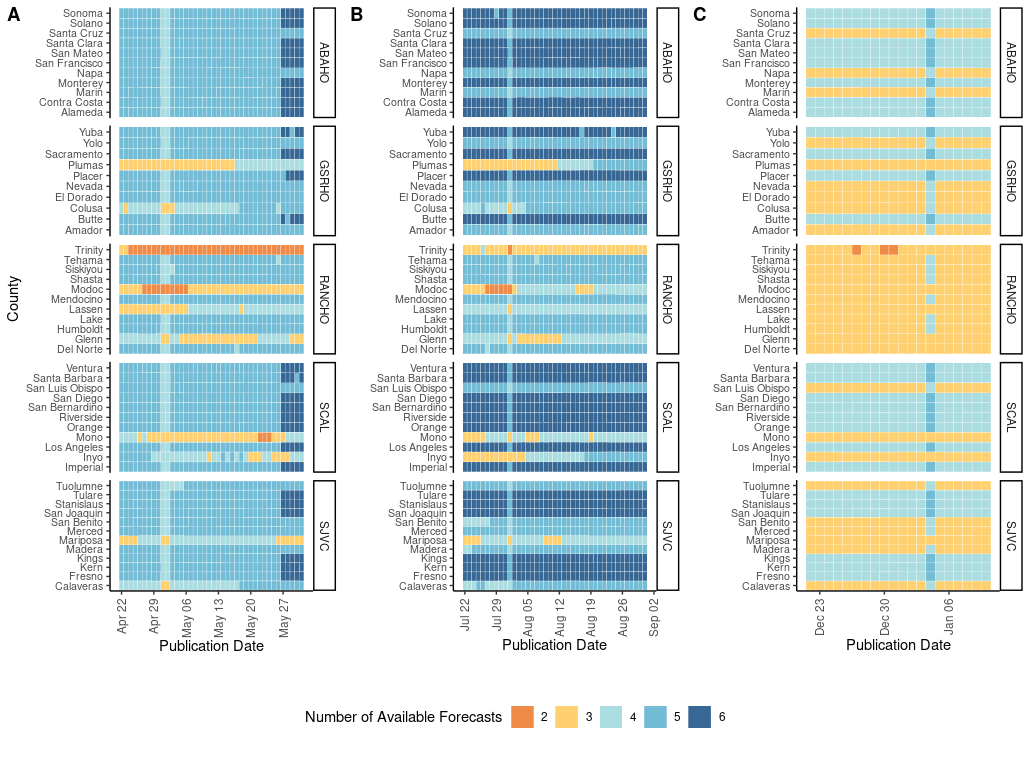

**Supplementary Figure 15. Number of available forecasts for a given location and date for the (A) Alpha, (B) Delta, and (C) Omicron variant periods for 21-day MAE.**

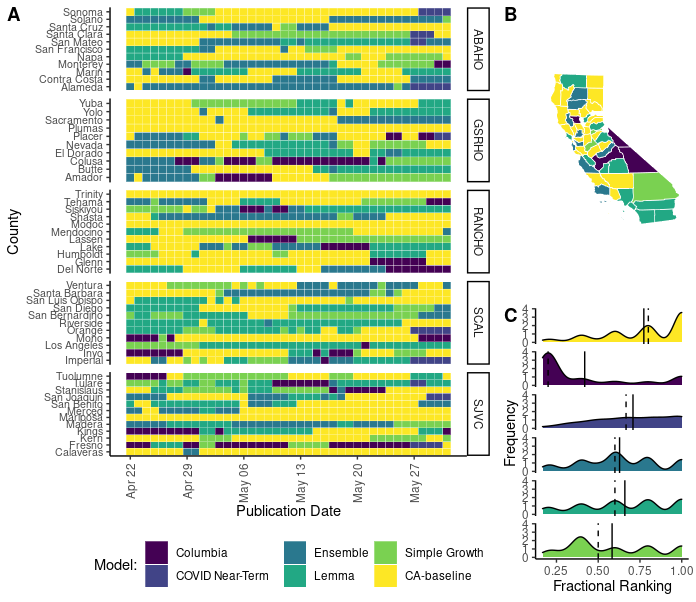

**Supplementary Figure 16. Forecasting accuracy results at the county level during the Alpha wave in California as measured by 21-day mean absolute error (MAE).** **(A)** Heat map of the best daily performing model for a given prediction date as measured by 21-day MAE. Each cell in the heat map corresponds to a standardized mean absolute error calculated for the day that a model forecast was published. Counties are grouped into panels by California health officer regions. **(B)** A summary map of California where the color of the county corresponds to the model with the highest sum of the standardized rank score for that period $({\Sigma sr}_{m,i,j})$. Note that by using the summation of the standardized ranking score models are penalized for lack of participation. **(C)** A density distribution of the standardized rank score $({sr}_{m,i,j})$ that depicts the median (dashed) and mean (solid) as vertical lines.

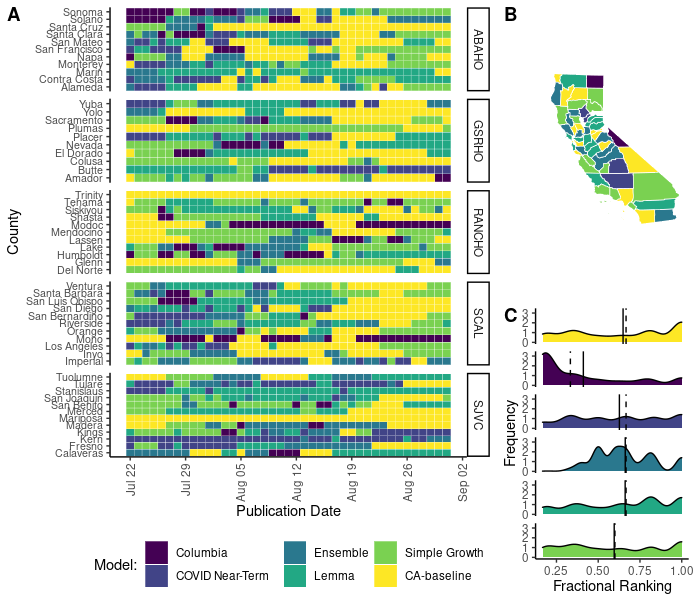

**Supplementary Figure 17. Forecasting accuracy results at the county level during the Delta wave in California as measured by 21-day mean absolute error (MAE).** **(A)** Heat map of the best daily performing model for a given prediction date as measured by 21-day MAE. Each cell in the heat map corresponds to a standardized mean absolute error calculated for the day that a model forecast was published. Counties are grouped into panels by California health officer regions. **(B)** A summary map of California where the color of the county corresponds to the model with the highest sum of the standardized rank score for that period $({\Sigma sr}_{m,i,j})$. Note that by using the summation of the standardized ranking score models are penalized for lack of participation. **(C)** A density distribution of the standardized rank score $({sr}_{m,i,j})$ that depicts the median (dashed) and mean (solid) as vertical lines.

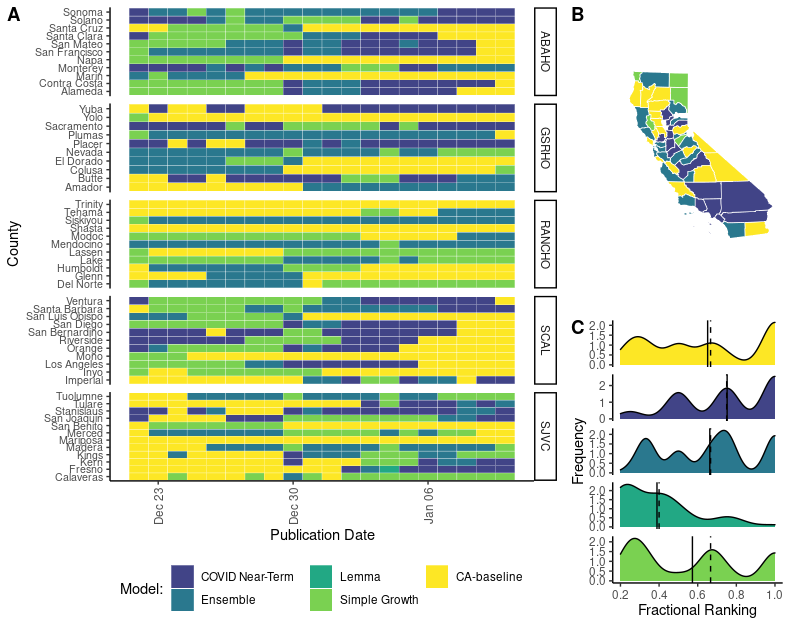

**Supplementary Figure 18. Forecasting accuracy results at the county level during the Omicron wave in California as measured by 21-day mean absolute error (MAE). (A)** Heat map of the best daily performing model for a given prediction date as measured by 21-day MAE. Each cell in the heat map corresponds to a standardized mean absolute error calculated for the day that a model forecast was published. Counties are grouped into panels by California health officer regions. **(B)** A summary map of California where the color of the county corresponds to the model with the highest sum of the standardized rank score for that period $({\Sigma sr}_{m,i,j})$. Note that by using the summation of the standardized ranking score models are penalized for lack of participation. **(C)** A density distribution of the standardized rank score $({sr}_{m,i,j})$ that depicts the median (dashed) and mean (solid) as vertical lines.

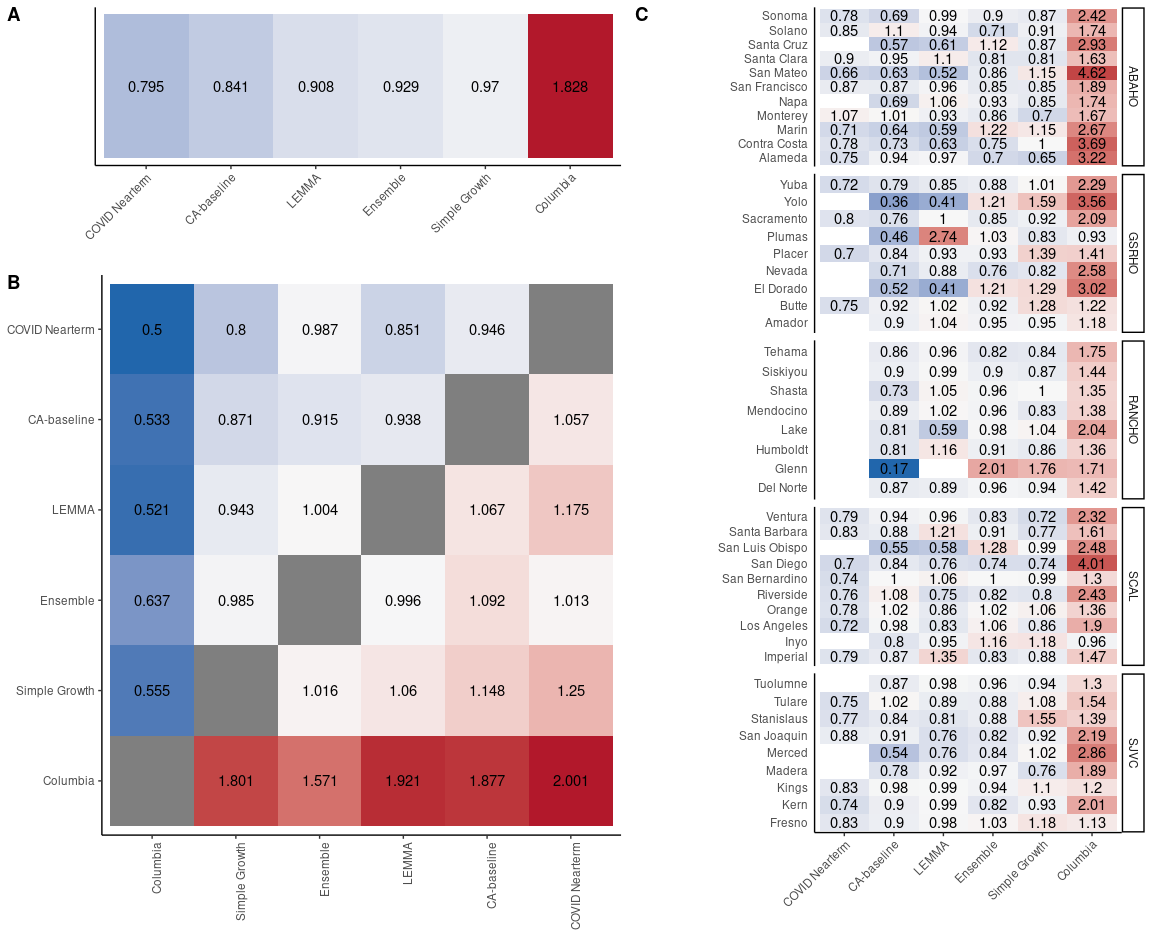

**Supplementary Figure 19.** **Pairwise tournament median rankings of models during the whole analysis period (January 1, 2021-February 2, 2022) for 21-day MAE.** **(A)** Overall median ranking across all locations and observation dates. **(B)** Median pairwise ranking comparing each model *m* to all available models *M*. Grid is symmetrical, so Model 1: Model 2 = 1/(Model 2:Model 1). **(C)** Overall median rankings for all available observation dates disaggregated by county.

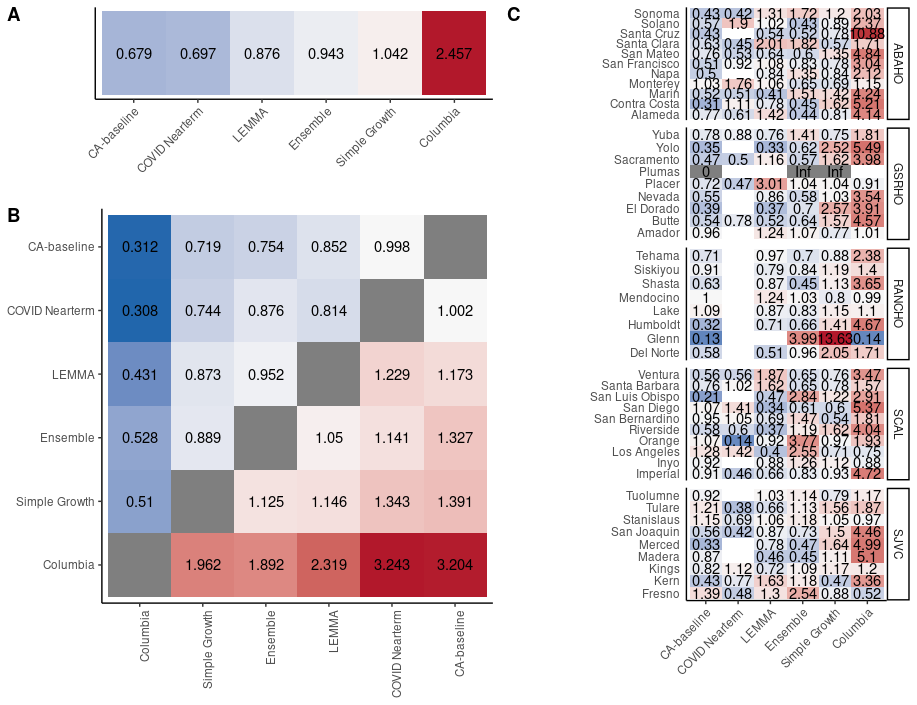

**Supplementary Figure 20.** **Pairwise tournament median rankings of models during the Alpha period for 21-day MAE. (A)** Overall median ranking across all locations and observation dates. **(B)** Median pairwise ranking comparing each model *m* to all available models *M*. Grid is symmetrical, so Model 1: Model 2 = 1/(Model 2:Model 1). **(C)** Overall median rankings for all available observation dates disaggregated by county.

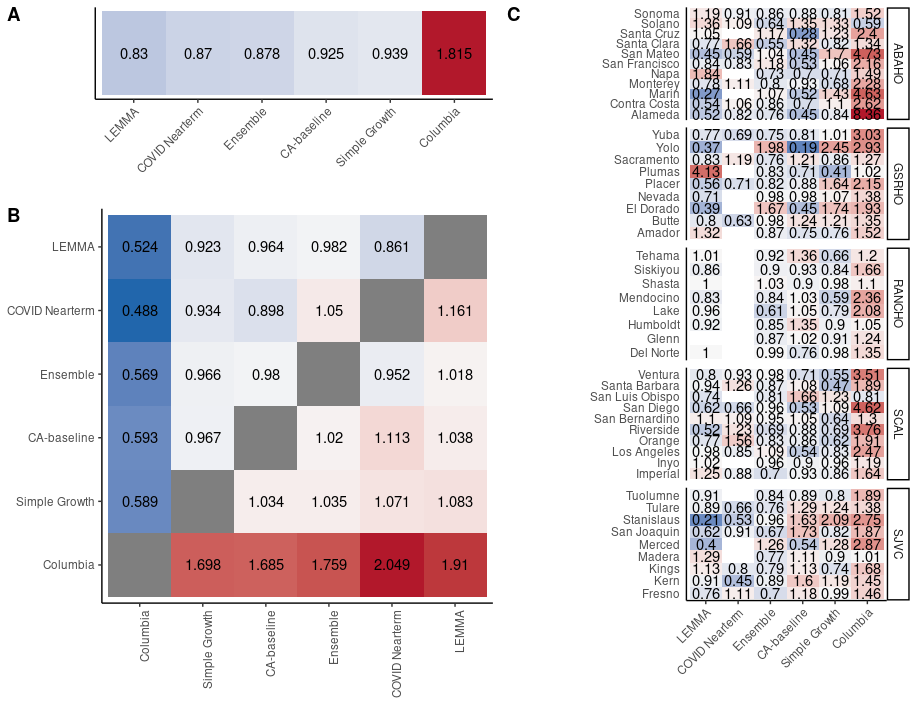

**Supplementary Figure 21.** **Pairwise tournament median rankings of models during the Delta period for 21-day MAE.** **(A)** Overall median ranking across all locations and observation dates. **(B)** Median pairwise ranking comparing each model *m* to all available models *M*. Grid is symmetrical, so Model 1: Model 2 = 1/(Model 2:Model 1). **(C)** Overall median rankings for all available observation dates disaggregated by county.

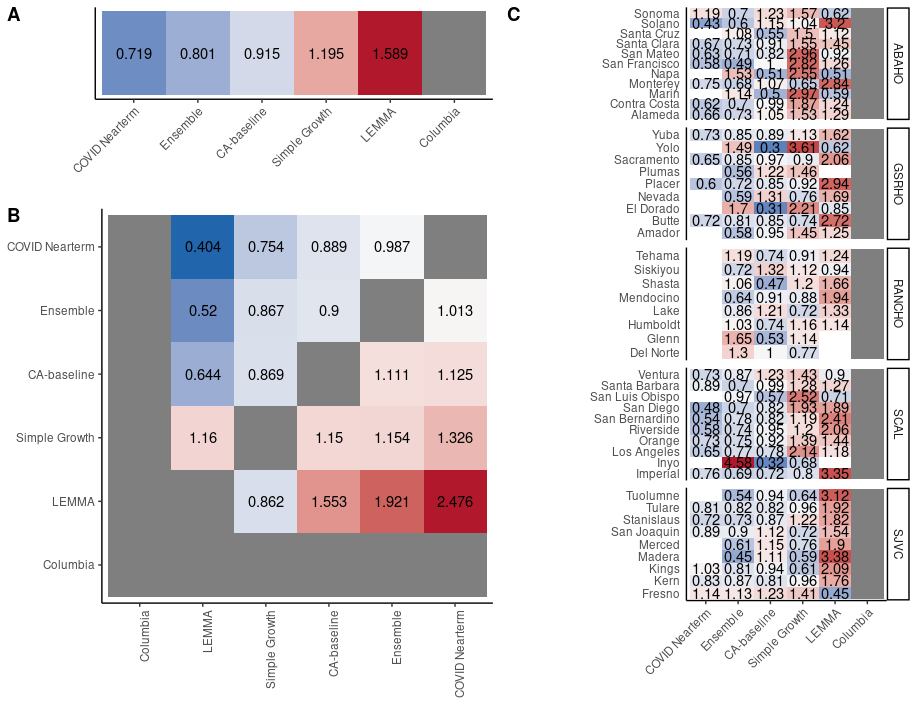

**Supplementary Figure 22. Pairwise tournament median rankings of models during the Omicron period for 21-day MAE. (A)** Overall median ranking across all locations and observation dates. **(B)** Median pairwise ranking comparing each model *m* to all available models *M*. Grid is symmetrical, so Model 1: Model 2 = 1/(Model 2:Model 1). **(C)** Overall median rankings for all available observation dates disaggregated by county.

### Random forest variable importance

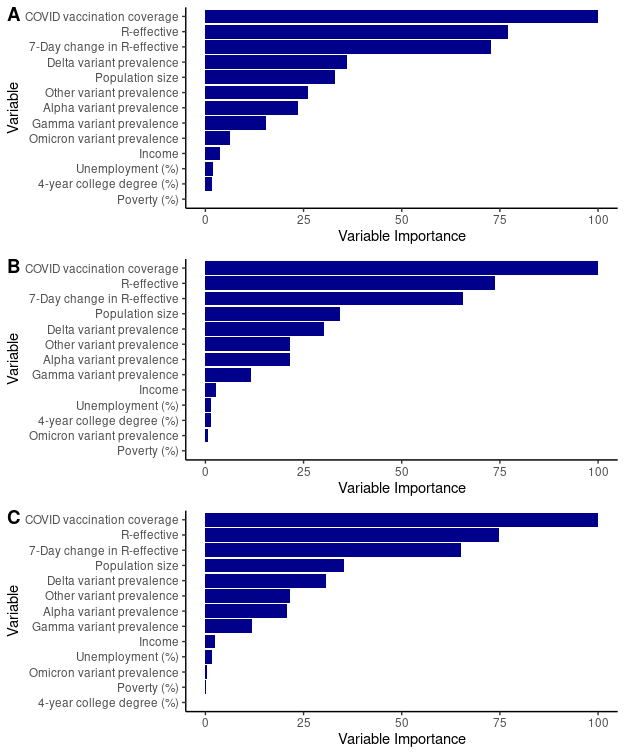
 **Supplementary Figure 23. Variable importance values from random forest classification analysis for (A) 7 day, (B) 14-day, and (C) 21-day MAE.**

### MAE vs. county size

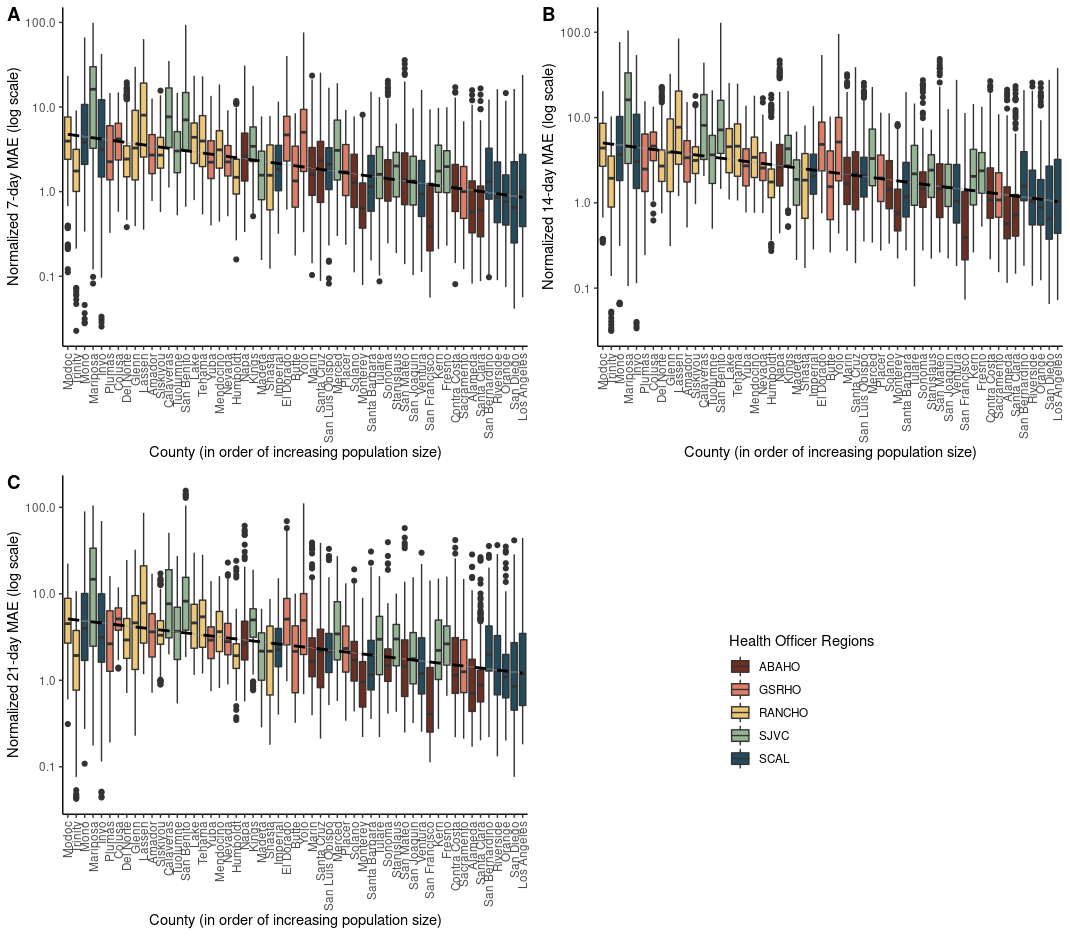

**Supplementary Figure 24. Boxplots of normalized MAE distributions by county for (A) 7-day, (B) 14-day, and (C) 21-day MAE for the ensemble model.** MAE is normalized by median county hospital capacity. Counties are situated on the x-axis in order of increasing population size. Counties are color-coded based on their health officer region membership. Distribution of MAE scores are from the entire analysis period: February 1, 2021- February 1, 2022.
